# Association of the timing and type of acute symptomatic seizures with post-stroke epilepsy and mortality

**DOI:** 10.1101/2024.12.02.24318052

**Authors:** Kai Michael Schubert, Dominik Zieglgänsberger, Giulio Bicciato, Laura Abraira, Estevo Santamarina, José Álvarez-Sabín, Carolina Ferreira-Atuesta, Mira Katan, Lucia Sinka, Robert Terziev, Nico Döhler, Barbara Erdélyi-Canavese, Ansgar Felbecker, Philip Siebel, Michael Winklehner, Tim J von Oertzen, Judith N. Wagner, Gian Luigi Gigli, Annacarmen Nilo, Francesco Janes, Giovanni Merlino, Mariarosaria Valente, María Paula Zafra-Sierra, Luis Carlos Mayor-Romero, Julian Conrad, Stefan Evers, Matias Alet, Kazuki Fukuma, Masafumi Ihara, Benjamin Landau, Piergiorgio Lochner, Frauke Roell, Francesco Brigo, Carla Bentes, Ana Rita Peralta, Teresa Pinho e Melo, Mark R Keezer, John S Duncan, Josemir W Sander, Barbara Tettenborn, Matthias J Koepp, Marian Galovic

## Abstract

**Background:** Acute symptomatic seizures (ASyS) increase the risk of epilepsy and mortality after a stroke. The impact of the timing and type of ASyS remains unclear.

**Methods:** This multicenter cohort study included data from nine centers between 2002 and 2018, with final analysis in February 2024. The study included 4,552 adults (2,005 female; median age 73 years) with ischemic stroke and no seizure history. We examined ASyS occurring within seven days after stroke. Main outcomes were all-cause mortality and epilepsy. Validation in three separate cohorts included 74 adults with ASyS.

**Results:** The ten-year risk of post-stroke epilepsy ranged from 41% to 94%, and mortality from 36% to 100%, depending on ASyS type and timing. ASyS on stroke onset day had a higher epilepsy risk (aHR 2.3, 95% CI 1.3-4.0, p=0.003) compared to later ASyS. Status epilepticus had the highest epilepsy risk (aHR 9.6, 95% CI 3.5-26.7, p<0.001), followed by focal to bilateral tonic-clonic seizures (aHR 3.4, 95% CI 1.9-6.3, p<0.001). Mortality was higher in those with ASyS presenting as focal to bilateral tonic-clonic seizures on day 0 (aHR 2.8, 95% CI 1.4-5.6, p=0.004) and status epilepticus (aHR 14.2, 95% CI 3.5-58.8, p<0.001). The novel SeLECT-ASyS model, available as an app, outperformed a previous model in the derivation cohort (concordance statistic 0.68 vs. 0.58, p=0.02) and in the validation cohort (0.70 vs. 0.50, p=0.18).

**Conclusions:** ASyS timing and type significantly affect epilepsy and mortality risk after stroke, improving epilepsy prediction and guiding patient counseling.

## Introduction

Stroke is a major cause of epilepsy in older adults, contributing to over half of new-onset epilepsy cases in individuals aged 65 and above.^1^ Post-stroke seizures are associated with an increased risk of mortality, poor functional outcome, disability, and dementia.^2,3^

Post-stroke seizures are categorized into acute symptomatic (ASyS), occurring within the first 7 days after a stroke, and remote symptomatic (RSyS), which are unprovoked seizures occurring later.^4^ ASyS are considered provoked and do not qualify as epilepsy. In contrast, a single or multiple RSyS following ischemic stroke fulfill the International League Against Epilepsy (ILAE) practical definition of epilepsy due to a heightened, greater than 60% recurrence risk of seizures.^5^

ASyS are a major risk factor for epilepsy and mortality following ischemic stroke.^3,6^ Recent research underscores the heterogeneity among ASyS, suggesting certain subtypes confer higher risks than others.^7^ We have shown that ASyS presenting as status epilepticus carry a markedly elevated risk of epilepsy and mortality compared to short ASyS after ischemic stroke.^3^ However, there remains a knowledge gap regarding other characteristics of ASyS that may be associated with an increased risk of seizures or unfavorable outcomes.

We hypothesized that the timing and type of a short ASyS influence the risk of post-stroke epilepsy and mortality. We assessed this hypothesis using data from a large multicenter registry of post-stroke seizures. We implemented this knowledge in a novel prognostic model that improves the prediction of epilepsy following ASyS after ischemic stroke.

## Methods

### Participants

We analyzed both a derivation and a validation cohort of participants. The derivation cohort was drawn from a multicenter registry established as part of the SeLECT study,^6^ consisting of nine international cohorts (n=4,552) of adults with neuroimaging-confirmed acute ischemic stroke. The validation cohort included participants with ASyS following ischemic stroke from three additional independent cohorts. We excluded individuals with transient ischemic attacks, a history of seizures or epilepsy, primary hemorrhagic stroke, re-infarction during follow-up, or potentially epileptogenic comorbidities (such as intracranial tumors, cerebral venous thrombosis, severe traumatic brain injury, or prior brain surgery). Detailed descriptions of the individual cohorts are provided in the Online Supplement.

Informed consent, obtained either in written or verbal form (four cohorts utilized written consent, while two cohorts employed both written and verbal consent), was acquired. In three cohorts consent requirements were waived by the regulatory authorities, as outlined in the online supplement.

### Definitions

According to ILAE recommendations, seizures were classified as ASyS (occurring within 7 days after stroke) or RSyS (spontaneous unprovoked seizures more than 7 days after stroke).^4^ The occurrence of a RSyS was categorized as poststroke epilepsy due to its high seizure recurrence risk, exceeding the 60% risk required for the ILAE pragmatic definition of epilepsy.^5,8^ Status epilepticus was classified according to the revised ILAE definition.^9^ Electrographic status epilepticus was defined according to Salzburg and revised ACNS criteria. ^10,11^ ASyS not qualifying as status epilepticus were defined as short ASyS and classified into subtypes (focal aware, focal with impaired awareness, focal to bilateral tonic clonic, or undetermined) based on the current ILAE nomenclature.^12^ We dichotomized the timing of ASyS into those occurring on the same day as stroke onset (day 0) versus those occurring later because the majority of ASyS occurred on day 0. In individuals with multiple ASyS, we only considered the first reported ASyS. Further definitions are detailed in the eMethods in the Online Supplement.

### Statistical analysis

Firstly, we used multivariable Cox proportional hazards regression to assess the relationship between the type and timing of ASyS and the time to post-stroke epilepsy or death, while adjusting for co-variates (age, sex, National Institutes of Health Stroke Scale [NIHSS] score at admission, cortical involvement, involvement of the middle cerebral artery [MCA] territory, stroke cause, reperfusion treatment, and antiseizure medication [ASM] treatment after acute symptomatic seizure). Cases were censored at the time of death, first RSyS, or last follow-up. Adjusted risk estimates for post-stroke epilepsy or death were obtained from these multivariable Cox regression models.

Secondly, we compared the performance of a previously described prognostic model for post-stroke epilepsy (SeLECT_2.0_)^3^ in stroke survivors with vs. without ASyS using the concordance (C) statistic. We also compared the observed risk of seizures in those with vs. without ASyS having a similar SeLECT_2.0_ score strata (3 to 4 points and 5 to 6 points) using Kaplan Meier estimate plots and logrank tests.

Next, we developed a novel prognostic model specifically for stroke survivors with ASyS with the goal to improve the prediction of post-stroke epilepsy in this particularly vulnerable population. Least Absolute Shrinkage and Selection Operator (LASSO) Cox-regression was employed initially for variable selection, utilizing a penalty function to refine the model.^13^ The regularization parameter lambda in the LASSO regression was selected using k-fold cross-validation (k=10). Following the LASSO selection, we employed Wald stepwise backward regression as an additional method to identify significant variables and compare them with those selected by LASSO. To account for death as a competing risk, we also employed competing risk regression based on Fine and Gray’s subdistribution hazard model, which allowed us to calculate the cumulative incidence function for late seizures (see **Supplemental Table 4**). We assigned integer values to the retained variables based on their adjusted hazards ratios (aHR, see **Supplemental Table 5**) to calculate a clinical risk score for each study individual.

To evaluate the novel model’s discrimination, i.e. the ability to distinguish between high and low risk cases, we estimated the C statistic (95% CI). Recognizing that prognostic models derived from multivariable regression can exhibit optimism and potentially overestimate predictions when applied to new patient cohorts,^14^ we introduced a shrinkage factor. This factor was estimated through 1000 bootstrapped random samples to adjust the C statistic for overoptimism, a technique previously employed to enhance model generalizability.^15,16^ We also assessed model calibration, i.e. the agreement between predicted and observed risks, using calibration plots (see **Supplemental Figure 5**). Perfect calibration is represented by a 45° diagonal line, whereas relevant deviation above or below this line reflects under- or overprediction. We used a leave-one-cohort-out strategy for cross-validation of model performance.

Lastly, we computed the change of occurrence of a seizure in the next year (COSY), a parameter that may be relevant for assessing the fitness to drive in people with seizures, using the standard statistical definition of conditional risks^17^ as detailed previously (Schubert et al. 2023) (see **Supplemental Figure 2**). Previously proposed COSY thresholds are < 20-40% for private driving and < 2% for professional driving,^18,19,20^ although these may differ based on local regulations.

All analyses were conducted using R statistical software version 4.0.3 and SPSS version 26 (IBM) and followed the STROBE guidelines.

## Results

The study included 4,552 individuals from nine centers. Their baseline characteristics are shown in **Supplemental Table 1**. ASyS occurred in 5% (n=233) of participants. The first ASyS presented on the same day as the stroke (day 0) in 55% (n=127, **Figure 1A** and **Supplemental Table 2**). The frequency of ASyS stratified by type and timing is shown in **Figure 1B**.

**Figure 1:**
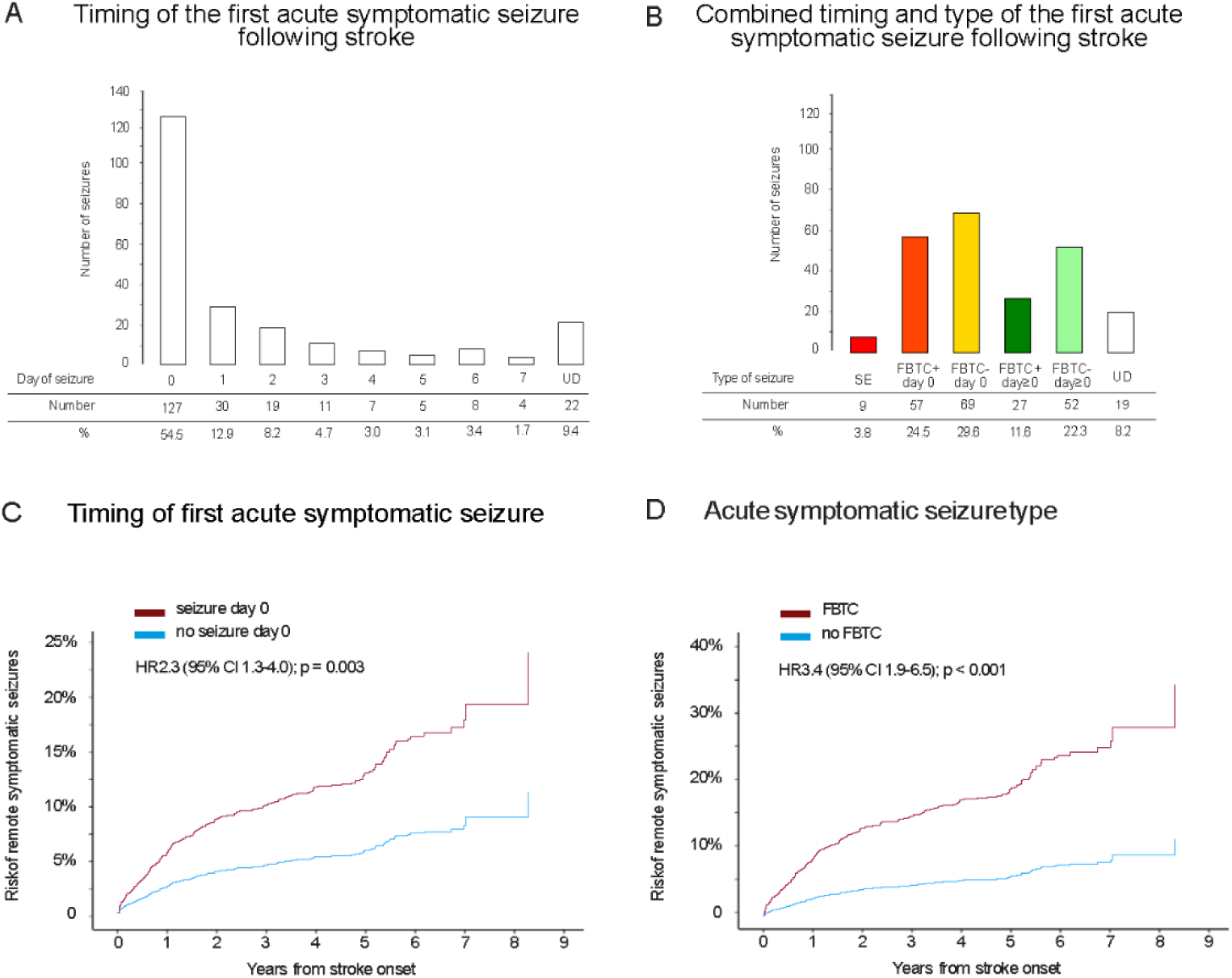
Characterization of acute symptomatic seizure timing and type. Acute symptomatic seizure, ASyS; UD, undetermined/unknown seizure timing. Panel A and B: Distribution of the timing (Panel A) and stratification by type and timing (Panel B) of the first acute symptomatic seizure occurring after ischemic stroke. Color code in Panel B adapted to the stratification of ASyS in Figure 3. Panels C and D: Kaplan Meier estimates (n = 4552) of the time to post-stroke epilepsy stratified by acute symptomatic seizure timing (day 0 vs. day ≥ 1; Panel B) and type (FBTCS vs. other short seizure type; Panel C).

### Risk of post-stroke epilepsy

ASyS on day 0 had a higher risk of post-stroke epilepsy (aHR 2.3, 95% CI 1.3-4.0, p=0.003, **Figure 1C**, **Table 1**) compared to ASyS occurring later after stroke. Other ASyS timing cut-offs did not yield relevant differences in the risk of post-stroke epilepsy (**Supplemental Figure 3**). Regarding seizure types, ASyS presenting as status epilepticus had the highest risk of post-stroke epilepsy (aHR 9.6, 95% CI 3.5-26.7, p<0.001), followed by FBTCS (aHR 3.4, 95% CI 1.9-6.3 p<0.001, **Figure 1D**, **Table 1**) compared to other ASyS types (**Supplemental Figure 4**). The full results of the multivariable model and other variables independently associated with post-stroke epilepsy (stroke severity, location, and etiology) are shown in **Table 1**.

**Table 1:** Multivariable Cox regression model of time to first remote symptomatic seizure.

| Variable | aHR (95% CI) | P value |
| --- | --- | --- |
| Demographics |  |  |
| Age (*per 10 years*) | 0.9 (0.9-1.0) | 0.07 |
| Male sex | 1.0 (0.8-1.3) | 0.91 |
| Stroke severity at admission |  |  |
| NIHSS 4-10 | **1.7 (1.2-2.5)** | **<0.001** |
| NIHSS $\geq 11$ | **4.2 (3.0-5.9)** | **<0.001** |
| Stroke location |  |  |
| Middle cerebral artery territory involvement | **1.5 (1.2-2.1)** | **0.02** |
| Cortical involvement | **2.2 (1.7-2.9)** | **<0.001** |
| Stroke cause |  |  |
| Small-vessel occlusion | 1.0 (0.6-1.6) | 0.90 |
| Larger-artery atherosclerosis | **1.6 (1.2-2.1)** | **0.003** |
| Cardioembolic | 0.9 (0.7-1.2) | 0.51 |
| Treatment |  |  |
| Acute reperfusion treatment | 0.8 (0.6-1.0) | 0.05 |
| Acute symptomatic seizure type |  |  |
| Seizure occurring on day 0 | **2.3 (1.3-4.0)** | **0.003** |
| Focal seizure with impaired awareness | 1.6 (0.7-3.9) | 0.28 |
| Focal to bilateral tonic-clonic seizure | **3.4 (1.9-6.3)** | **<0.001** |
| Status epilepticus | **9.6 (3.5-26.7)** | **<0.001** |
| Undetermined or unknown seizure type | 1.8 (0.7-4.1) | 0.20 |

Data analyzed using a Cox proportional hazards model in the derivation cohort (n=4552).
Dependent variable was time to first remote seizure.
aHR, adjusted hazard ratio; NIHSS, National Institutes of Health Stroke Scale; ASM, anti-seizure medication; ASyS, acute symptomatic seizure.

Stroke survivors with a non-FBTC short acute symptomatic seizure occurring on day 1 or later after stroke had a 41% risk of developing post-stroke epilepsy 10 years after stroke, compared to a 69% risk in those having a FBTC short acute symptomatic seizure on day 0 after stroke and a 94% risk in those with acute symptomatic status epilepticus (**Figure 3A**, **Supplemental Table 2**). The 10-year risk of post-stroke epilepsy was 13% in stroke survivors without acute symptomatic seizures.

### Mortality

A higher risk of all-cause mortality was observed in those with ASyS presenting as FBTCS on day 0 (aHR 2.8, 95% CI 1.4-5.6, p=0.004) and those with acute symptomatic status epilepticus (aHR 14.2, 95% CI 3.5-58.8, p<0.001). Other subtypes of acute symptomatic seizures were not associated with mortality (**Table 2**).

**Table 2:** Multivariable Cox regression model of time to death.

| Variable | aHR (95% CI) | P value |
| --- | --- | --- |
| Demographics |  |  |
| Age (*per 10 years*) | **2.2 (2.0-2.4)** | **<0.001** |
| Male sex | **1.2 (1.0-1.4)** | **0.05** |
| Stroke severity at admission |  |  |
| NIHSS 4-10 | **1.7 (1.3-2.1)** | **<0.001** |
| NIHSS $\geq 11$ | **2.4 (1.9-3.1)** | **<0.001** |
| Stroke location |  |  |
| Middle cerebral artery territory involvement | 0.9 (0.8-1.2) | 0.64 |
| Cortical involvement | 1.0 (0.9-1.3) | 0.71 |
| Stroke cause |  |  |
| Small-vessel occlusion | **0.6 (0.4-0.8)** | **<0.001** |
| Larger-artery atherosclerosis | **0.7 (0.5-0.9)** | **0.02** |
| Cardioembolic | 0.8 (0.7-1.0) | 0.07 |
| Treatment |  |  |
| Acute reperfusion treatment | **0.8 (0.7-1.0)** | **0.04** |
| ASM treatment after ASyS | **0.5 (0.3-0.9)** | **0.03** |
| Acute symptomatic seizure type |  |  |
| FBTC-, day > 0 | 1.8 (0.8-4.0) | 0.18 |
| FBTC+, day > 0 | 1.2 (0.4-3.7) | 0.81 |
| FBTC-, day 0 | 1.4 (0.7-2.6) | 0.32 |
| FBTC+, day 0 | **2.8 (1.4-5.6)** | **0.004** |
| Status epilepticus | **14.2 (3.5-58.8)** | **<0.001** |

Data analyzed using a Cox proportional hazards model in the derivation cohort (n=4552).
Dependent variable was time to death of any cause.
aHR, adjusted hazard ratio; NIHSS, National Institutes of Health Stroke Scale; ASM, anti-seizure medication; ASyS, acute symptomatic seizure; FBTC+, focal to bilateral tonic-clonic short seizure; FBTC-, other type of short seizure; day 0, same day as stroke onset.

Individuals with a stroke and a non-FBTC short acute symptomatic seizure occurring on day 1 or later after stroke had a 49% risk of all-cause mortality 10 years after stroke, compared to a 66% risk in those having a FBTC short acute symptomatic seizure on day 0 after stroke and a 100% risk in those with acute symptomatic status epilepticus (**Figure 3B**). The overall 10-year risk of all-cause mortality was 32% in those without acute symptomatic seizures.

### Prognostic modelling

We observed that the performance of a previously published prognostic model predicting the risk of post-stroke epilepsy (SeLECT_2.0_) was low in individuals with ASyS (C statistic 0.58, 95%-CI 0.49-0.67, n=233) compared to a better performance in the overall cohort (C statistic 0.75, 95%-CI 0.72-0.78, n=4,552). The observed risk of post-stroke epilepsy in stroke survivors with ASyS was higher compared to those without ASyS who had a similar SeLECT_2.0_ score (**Figure 2**; SeLECT_2.0_ score 3 to 4, p<0.001; SeLECT_2.0_ score 5 to 6, p=0.10).

**Figure 2:**
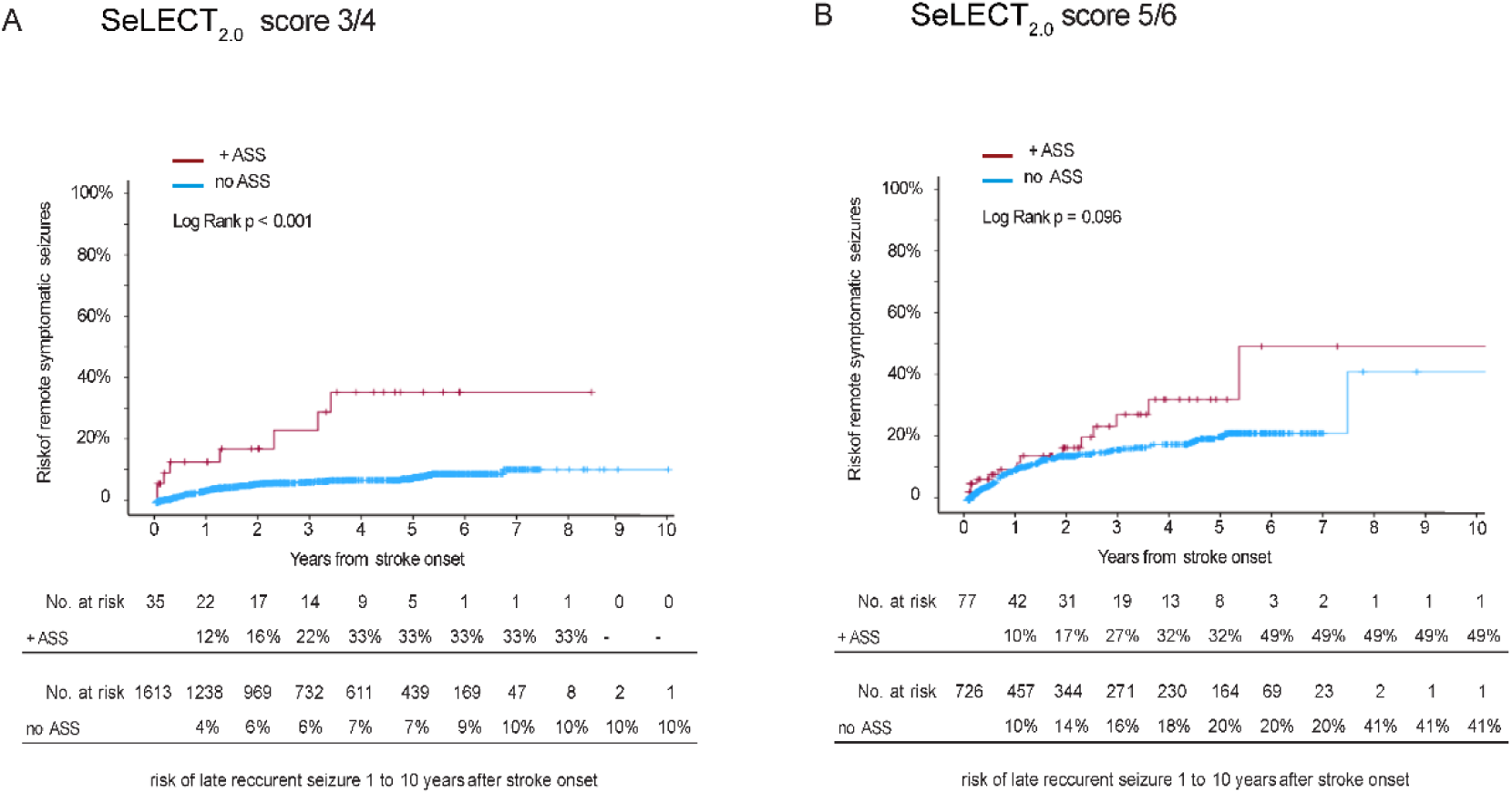
Risk of epilepsy in individuals with or without acute symptomatic seizures having a similar SeLECT_2.0_ score. Kaplan-Meier estimates (n = 4552) of the time to post-stroke epilepsy in those with a SeLECT_2.0_ score of 3 to 4 (Panel A) or 5 to 6 (Panel B) points. Separate curves are shown for individuals with (red) or without (blue) acute symptomatic seizures. Those with a SeLECT_2.0_ score of 3 to 4 who suffered acute symptomatic seizures had a higher risk of post-stroke epilepsy (higher risk of remote symptomatic seizures) compared to those without acute symptomatic seizures (p<0.001). There was a similar but non-significant trend in individuals with a SeLECT_2.0_ score of 5 to 6 who had acute symptomatic seizures compared to those without (p=0.096).

**Figure 3:**
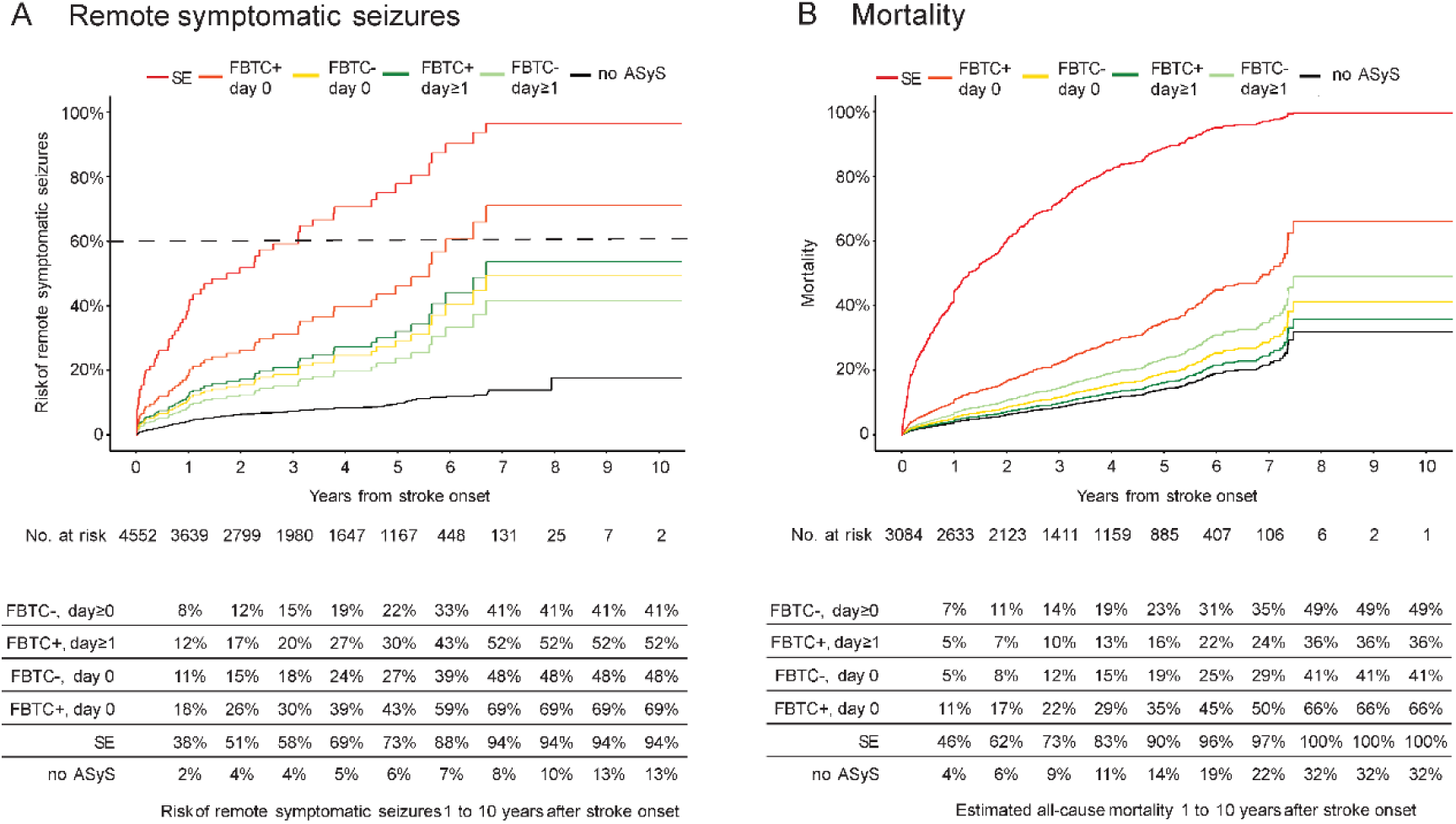
Risk of Post-Stroke Epilepsy or Mortality Following ASyS After Stroke. Data according to the type and timing of ASyS and association with RSySs (n = 4552, Panel A) and mortality (n= 3084, Panel B). The tables below each graph display the Kaplan-Meier estimates of the risk of RSySs 1 to 10 years after index stroke according to the type of ASyS. All results were obtained after adjusting for covariates (age, sex, National Institutes of Health Stroke Scale score at admission, cortical involvement, involvement of the middle cerebral artery territory, stroke cause, reperfusion treatment, and antiseizure medication treatment after ASyS). The dotted line denotes the 60% cutoff for the risk of unprovoked seizures used in the International League Against Epilepsy (ILAE) practical clinical definition of epilepsy. FBTC indicates focal to bilateral tonic-clonic seizure; no ASyS, no ASyS; SE, status epilepticus; and UD day, undetermined or unknown seizure day.

Thus, the SeLECT_2.0_ model may not adequately capture the risk of epilepsy in those with ASyS. To overcome these limitations, we aimed to develop a model specifically for those having ASyS after stroke and to implement the above findings on the risk of epilepsy according to ASyS type and timing.

We selected predictors using stepwise backward-elimination of a Cox regression model in stroke survivors with ASyS (**Supplemental Table 4**). The variables retained in the final model were timing and type of first ASyS, large-vessel atherosclerotic stroke etiology stratified by sex and stroke involving the cerebral cortex. The calculation of the new model, termed SeLECT-ASyS and ranging from 0 to 7 points, is shown in **Table 3**. The SeLECT-ASyS model had a better discrimination for time to post-stroke epilepsy compared to the original SeLECT_2.0_ model (C statistic 0.68 [95%-CI 0.61-0.76] vs. 0.58 [95%-CI 0.49-0.67], p=0.02) in stroke survivors with ASyS. SeLECT-ASyS demonstrated better, near-optimal calibration for long-term outcomes compared to the less optimal calibration of the SeLECT_2.0_ model in those with ASyS, although the calibration curves indicated less accuracy for predicting the 1-year occurrence of post-stroke epilepsy and lacked data for lower probability outcomes (**Supplemental Figure 5**). We cross-validated the results using a leave-one-cohort-out strategy (**Supplemental Table 6**). To further support our findings, we evaluated a validation cohort of 74 adults with ASyS following ischemic stroke who met the eligibility criteria (Switzerland, n=32; Argentina, n=23; Japan, n=19). Baseline characteristics of these individuals are presented in **Supplemental Table 7**. In this validation cohorts, the SeLECT-ASyS model demonstrated superior discrimination with a C-statistic of 0.70 (95%-CI 0.57-0.83) compared to the SeLECT 2.0 model (C-statistic 0.59 [95%-CI 0.44-0.75], p=0.18), indicating better predictive accuracy for RSyS (**Supplemental Figure 6)**.

**Table 3:** Calculation of the SeLECT-ASyS score.

| Variable | SeLECT-ASyS<br>score (points) |
| --- | --- |
| Stroke location |  |
| Cortical involvement | 1 |
| Stroke etiology and sex |  |
| Large-artery atherosclerosis, female | 1 |
| Large-artery atherosclerosis, male | 2 |
| Acute symptomatic seizures type |  |
| FBTC-, day $\geq 1$ | 0 |
| FBTC+, day $\geq 1$ | 1 |
| FBTC-, day 0 | 1 |
| FBTC+, day 0 | 2 |
| Status epilepticus | 4 |

To calculate an individual's SeLECT-ASyS score, the points associated with each predictor can be added to obtain the total risk score. The total score ranges from 0 to 7 points. FBTC+, focal to bilateral tonic-clonic short seizure; FBTC-, other type of short seizure; day 0, same day as stroke onset.

The lowest score SeLECT-ASyS value (0 points) predicts a 26% risk (95% CI 9-39) of post-stroke epilepsy 10 years following stroke, compared with a 100% risk (95% CI 30-100) for the highest value (7 points, **Figure 4**). A comparison to the SeLECT_2.0_ values for patients without ASyS are shown in **Supplemental Figure 1**. In addition, we updated the values for COSY (**Supplemental Figure 2**), a parameter that may be helpful for assessing the risks for safe driving. The new SeLECT-ASyS model was implemented in the “SeLECT score” smartphone applications available for iOS and Android and in the web-based calculator.

**Figure 4:**
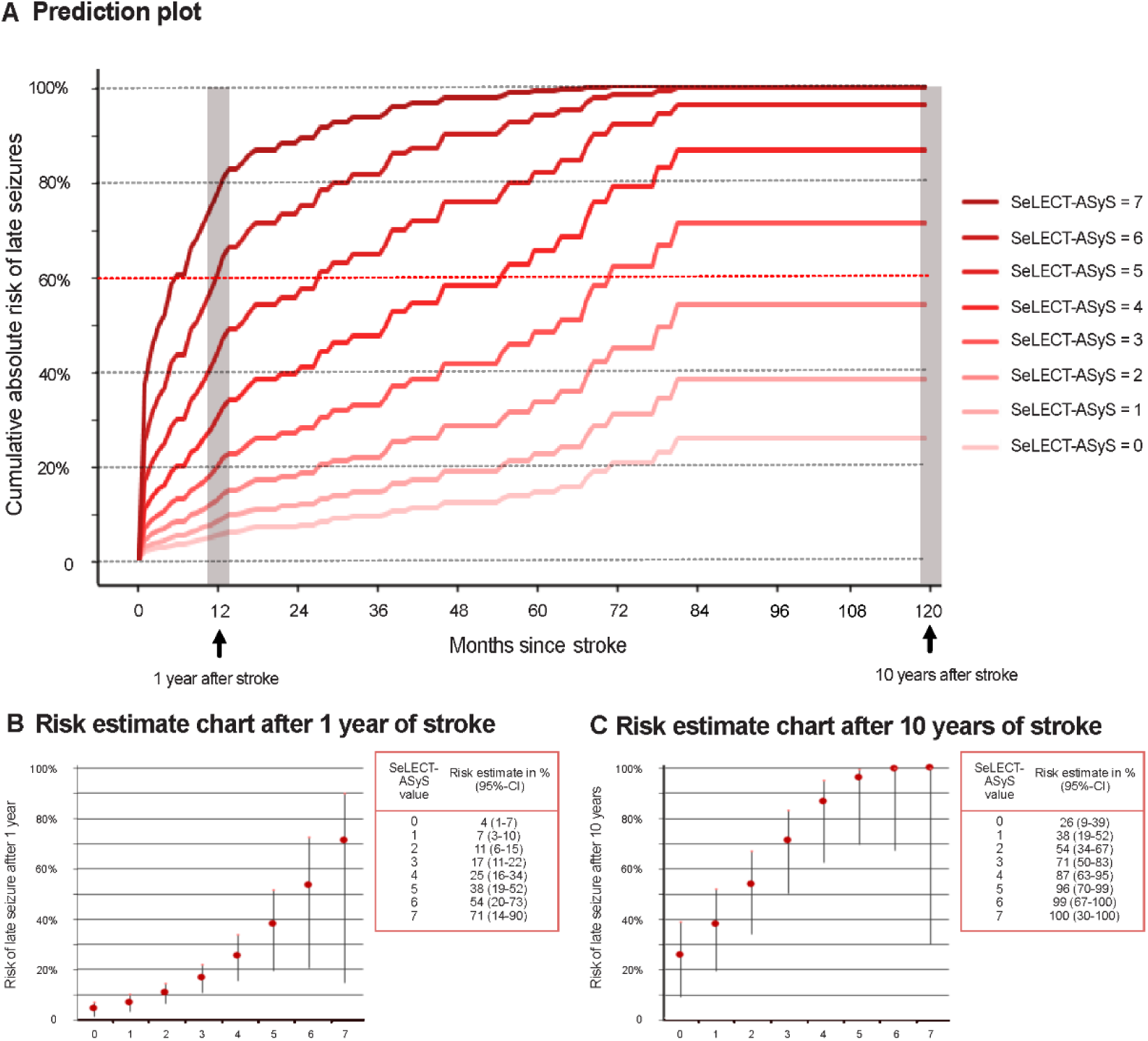
Predicted Risk of Post-Stroke Epilepsy According to a new prognostic model in stroke survivors with acute symptomatic seizures. Panel A shows the predicted risk of post-stroke epilepsy (unprovoked remote symptomatic seizures) 0-120 months after stroke. Each curve represents the estimates for a SeLECT-ASyS value ranging from 0 to 6. Risk estimate charts of late seizures 1 year and 10 years after stroke according to SeLECT-ASyS score values are displayed in panels B and C, respectively. Vertical lines indicate 95% CIs.

### Case Example

As an example, we considered a 65-year-old male patient admitted to the hospital because of acute ischemic stroke of atherosclerotic etiology affecting the posterior cerebral artery territory and not involving the cortex with 3 points on the NIHSS at admission, who had a focal to bilateral tonic-clonic seizure on the first day of admission. The SeLECT_2.0_ score for this case is 4 points, predicting a 19% risk (95% CI 8-28) of post-stroke epilepsy within 10 years after stroke. In contrast, the new model developed specifically for those with acute symptomatic seizures (SeLECT-ASyS, **Table 3**) is 4 points, predicting 87% (95% CI 63-95) risk of post-stroke epilepsy within 10 years.

## Discussion

In this study, we investigated the influence of both timing and type of ASyS on the risk of epilepsy and mortality after ischemic stroke. Our findings reveal substantial heterogeneity among ASyS, indicating that events occurring on the day of stroke onset (day 0) and those manifesting as FBTCS or status epilepticus had a higher risk of developing post-stroke epilepsy. FBTCS on day 0 and acute symptomatic status epilepticus had a high, ≥ 69% risk of epilepsy and a ≥ 66% risk of mortality (**Figure 3**).

We also showed that a current state-of-the-art model for predicting post-stroke epilepsy (SeLECT_2.0_) underperformed in stroke survivors with ASyS and confirmed this finding in independent validation cohorts. We implemented our results on the role of timing and types of ASyS in a novel model (SeLECT-ASyS), tailored specifically for stroke survivors experiencing ASyS. This model accurately captures the elevated risks of post-stroke epilepsy in this subgroup and significantly outperforms the SeLECT_2.0_ model in those with ASyS.

We previously showed that acute symptomatic status epilepticus is a predictor of mortality and epilepsy after stroke^3^ and discussed the potential explanations for this observation. The estimated risks for epilepsy and mortality in the current study are slightly higher compared to our previous study^3^ because of the completion of long-term (60-month) follow-up in the second largest cohort in the registry (Switzerland (2)) that results in more robust estimates.

Building upon our prior research, we demonstrated in this study that ASyS presenting as FBTCS and occurring on day 0 also carry a higher risk of post-stroke epilepsy and mortality. There are several potential explanations for this finding.

Firstly, individuals experiencing ASyS very early after stroke, i.e. on day 0, and ASyS presenting as FBTCS may have a higher predisposition for generating seizures. This genetic or acquired vulnerability,^21^ conceptualized as a low “seizure threshold”,^22^ may predispose these individuals to an earlier onset of ASyS and the propagation of seizure activity across hemispheres. Such a low seizure threshold may also heighten the probability of subsequent RSyS during epileptogenesis.^22^

Secondly, the occurrence of ASyS on day 0 and a bilateral spread of seizures could be indicative of a more pronounced proepileptic impact of the stroke. Our previous research suggested that more severe strokes due to large artery atherosclerosis, affecting the MCA territory, are more likely to result in post-stroke epilepsy.^6^ But, the present study’s outcomes were adjusted for these variables, establishing independence from such factors. Other stroke characteristics warrant consideration. At a macroscopic level, the precise localization^23^ and connectivity of stroke may contribute to acute seizures and epileptogenesis. Strokes highly connected to the basal ganglia and cerebellum were found to be more likely to cause epilepsy.^24^ On a microscopic scale, cellular changes that are difficult to detect *in vivo* in humans, such as neurodegeneration, axonal and synaptic sprouting, blood-brain barrier damage, and inflammation,^21^ may promote acute seizures and epileptogenesis.

Thirdly, disparities between ASyS occurring on day 0 and those manifesting later may be associated with the time-dependent dynamics of pathological mechanisms following a stroke, such as neuroinflammatory changes and metabolic derangements.^25^ Consequently, ASyS occurring early might be triggered by distinct mechanisms compared to those occurring later, potentially resulting in variations in the risk of post-stroke epilepsy.

Lastly, ASyS may directly or indirectly contribute to epileptogenesis. While this concept has been consistently demonstrated in animal models of status epilepticus, it is less well established for brief seizures.^26^ Some experimental evidence suggests that brief convulsive seizures may also contribute to the process of epileptogenesis. ^26,27,28^

ASyS presenting as FBTCS on day 0 were independently associated with higher mortality after stroke. They may be a marker of significant macro- or microscopic neuronal disruption caused by stroke, as discussed above. Furthermore, convulsive seizures have been linked to excitotoxic damage in animal models, potentially contributing to poor outcomes.^29^ Convulsive seizures may also be associated with injuries, heightened metabolic demand, and aspiration leading to pneumonia, further impacting overall outcomes.

To translate these findings into clinical practice, we incorporated them into a novel prognostic model. Initially, we demonstrated that the current state-of-the-art prognostic model for post-stroke seizures, SeLECT_2.0,_ underestimates the risk of post-stroke epilepsy in the subset of stroke survivors with ASyS (**Figure 2**). Stroke survivors with ASyS represent a distinct group with unique predictors and a heightened risk of epilepsy compared to those without ASyS.^30^ Subsequently, we developed and validated a novel prognostic model, SeLECT-ASyS, which exhibited superior discrimination and calibration compared to SeLECT_2.0_ in the ASyS subgroup. We also performed internal validation using optimism correction through bootstrapping and cross-validation **(Supplemental Table 6).** Furthermore, we also confirmed SeLECT_2.0_’s underperformance in stroke survivors with ASyS in independent external validation cohorts. Consequently, SeLECT-ASyS emerges as the preferred model for prognostication in stroke survivors with ASyS.

The presented case example illustrates marked differences in estimated risks when utilizing SeLECT-ASyS as opposed to SeLECT_2.0_ for individuals with ASyS. These differences in risk are clinically meaningful and may have an impact on treatment considerations and the approach to follow-up in such cases. Risk estimates derived from the novel model consistently indicate moderate to high risks of post-stroke epilepsy following ASyS (**Figure 4**). These risks are realistic, as corroborated by the favorable calibration of the model (**Supplemental Figure 5**).

The most notable and practically relevant finding is a greater than 60% 10-year risk of epilepsy in stroke survivors with ASyS presenting as FBTCS on day 0 or status epilepticus (28% of all ASyS cases, **Figure 1B**) and those with SeLECT-ASyS scores ≥ 3 points. This risk level aligns with the ILAE practical definition of epilepsy.^5^ But, it is crucial to acknowledge that this definition requires at least one unprovoked seizure occurrence and is, hence, not fully met in cases that suffered only an ASyS. Nevertheless, some clinicians may consider counselling these high-risk cases as if they had epilepsy, potentially recommending primary preventive treatment^31,32^ or extended follow-up. It is important to note, however, that the efficacy of primary preventive treatment after stroke remains unproven. Two ongoing antiepileptogenesis trials in stroke survivors may offer novel insights into the treatment of cases at high risk of post-stroke epilepsy.^33,34^ If such treatments become available, the accurate prediction of risk of post-stroke epilepsy in those with ASyS will become central for the selection of candidates for antiepileptogenic medications.

Our study has several strengths. We assessed one of the largest multicenter cohorts of post-stroke seizures. We translated our findings into a user-friendly prognostic model accessible through both a smartphone and web applications. The novel model outperforms the current state-of-the-art model and its clinical significance was underscored through an illustrative case.

Our study has limitations. Firstly, despite the inclusion of nine international cohorts, enhanced statistical power and generalisability of our results could be further achieved by incorporating data from North America, Asia, or Africa. Secondly, the practical constraints of performing continuous EEG in the entire multicenter registry limited our evaluation to clinically apparent seizures, not considering electrographic seizures. Thirdly, data collected in the registry did not differentiate between seizures occurring immediately at stroke onset and those on the same day as the stroke. However, existing studies^35,36^ suggest that the majority of seizures on day 0 align with the immediate onset of the stroke. Fourthly, patients with ASyS may receive ASM treatment which may impact the risk of subsequent unprovoked seizures. To address this, we corrected all results for ASM treatment. Lastly, the SeLECT registry did not consistently collect data on the cause of death and long-term disability.

### Conclusions

We demonstrated varying mortality and epilepsy risks based on the type and timing of ASyS following stroke. We implemented these findings in a novel prognostic model (SeLECT-ASyS) that outperformed a previous model and is available as both smartphone and web application. The ten-year epilepsy risk in those with ASyS presenting as FBTCS on day 0 or status epilepticus and those with SeLECT-ASyS ≥ 3 points exceeded 60%, a cut-off used for the ILAE practical definition of epilepsy.^5^ These findings have the potential to inform counselling for stroke survivors with ASyS, particularly those with a high (>60%) risk for post-stroke epilepsy.

## Data Availability

All data produced in the present study are available upon reasonable request to the authors

## Acknowledgements

JSD, JWS and MJK are based at the NIHR University College London Hospitals Biomedical Research Centre, which the UK Department of Health sponsors. JWS’s current position is endowed by the UK Epilepsy Society and he receives research support from the Marvin Weil Epilepsy Research Fund and the Christelijke Vereniging voor de Verpleging van Lijders aan Epilepsie, Netherland.

## Authors Contributions

KMS, DZ and MG conceptualized and designed the study. ND, BEC, AF, PS, NS, GB, JS, CFA, LI, MK, LA, ES, JAS, MW, TJO, JNW, GLG, AS, FJ, GM, MV, GG, JC, SE, PL, FR, FB, CB, ARP, TPM, BT, MRK, JSD, JWS, BT, MJK, MG contributed to the acquisition and analysis of the data. KMS and MG contributed to the interpretation of the data. KMS and MG drafted the manuscript and figures. All co-authors revised the manuscript for intellectual content.

## References

1. Hauser WA, Annegers JF, Kurland LT. Incidence of epilepsy and unprovoked seizures in Rochester, Minnesota: 1935-1984. Epilepsia. May-Jun 1993;34(3):453-68. doi:10.1111/j.1528-1157.1993.tb02586.x

2. Misra S, Kasner SE, Dawson J, et al. Outcomes in Patients With Poststroke Seizures: A Systematic Review and Meta-Analysis. JAMA Neurol. Nov 1 2023;80(11):1155–1165. doi:10.1001/jamaneurol.2023.3240

3. Sinka L, Abraira L, Imbach LL, et al. Association of Mortality and Risk of Epilepsy With Type of Acute Symptomatic Seizure After Ischemic Stroke and an Updated Prognostic Model. JAMA Neurol. Jun 1 2023;80(6):605–613. doi:10.1001/jamaneurol.2023.0611

4. Beghi E, D’Alessandro R, Beretta S, et al. Incidence and predictors of acute symptomatic seizures after stroke. Neurology. Nov 15 2011;77(20):1785–93. doi:10.1212/WNL.0b013e3182364878

5. Fisher RS, Acevedo C, Arzimanoglou A, et al. ILAE official report: a practical clinical definition of epilepsy. Epilepsia. Apr 2014;55(4):475–82. doi:10.1111/epi.12550

6. Galovic M, Dohler N, Erdelyi-Canavese B, et al. Prediction of late seizures after ischaemic stroke with a novel prognostic model (the SeLECT score): a multivariable prediction model development and validation study. Lancet Neurol. Feb 2018;17(2):143–152. doi:10.1016/S1474-4422(17)30404-0

7. Herzig-Nichtweiss J, Salih F, Berning S, et al. Prognosis and management of acute symptomatic seizures: a prospective, multicenter, observational study. Ann Intensive Care. Sep 15 2023;13(1):85. doi:10.1186/s13613-023-01183-0

8. Hesdorffer DC, Benn EK, Cascino GD, Hauser WA. Is a first acute symptomatic seizure epilepsy? Mortality and risk for recurrent seizure. Epilepsia. May 2009;50(5):1102–8. doi:10.1111/j.1528-1167.2008.01945.x

9. Trinka E, Cock H, Hesdorffer D, et al. A definition and classification of status epilepticus--Report of the ILAE Task Force on Classification of Status Epilepticus. Epilepsia. Oct 2015;56(10):1515–23. doi:10.1111/epi.13121

10. Leitinger M, Trinka E, Gardella E, et al. Diagnostic accuracy of the Salzburg EEG criteria for non-convulsive status epilepticus: a retrospective study. Lancet Neurol. Sep 2016;15(10):1054–62. doi:10.1016/S1474-4422(16)30137-5

11. Hirsch LJ, Fong MWK, Leitinger M, et al. American Clinical Neurophysiology Society’s Standardized Critical Care EEG Terminology: 2021 Version. J Clin Neurophysiol. Jan 1 2021;38(1):1–29. doi:10.1097/WNP.0000000000000806

12. Fisher RS, Cross JH, French JA, et al. Operational classification of seizure types by the International League Against Epilepsy: Position Paper of the ILAE Commission for Classification and Terminology. Epilepsia. Apr 2017;58(4):522–530. doi:10.1111/epi.13670

13. Tibshirani R. The lasso method for variable selection in the Cox model. Stat Med. Feb 28 1997;16(4):385–95. doi:10.1002/(sici)1097-0258(19970228)16:4<385::aid-sim380>3.0.co;2-3

14. Harrell FE, Jr., Lee KL, Mark DB. Multivariable prognostic models: issues in developing models, evaluating assumptions and adequacy, and measuring and reducing errors. Stat Med. Feb 28 1996;15(4):361–87. doi:10.1002/(sici)1097-0258(19960229)15:4<361::Aid-sim168>3.0.Co;2-4

15. Schrag A, Siddiqui UF, Anastasiou Z, Weintraub D, Schott JM. Clinical variables and biomarkers in prediction of cognitive impairment in patients with newly diagnosed Parkinson’s disease: a cohort study. Lancet Neurol. Jan 2017;16(1):66–75. doi:10.1016/s1474-4422(16)30328-3

16. Backes D, Rinkel GJE, Greving JP, et al. ELAPSS score for prediction of risk of growth of unruptured intracranial aneurysms. Neurology. Apr 25 2017;88(17):1600–1606. doi:10.1212/wnl.0000000000003865

17. Zabor EC, Gonen M, Chapman PB, Panageas KS. Dynamic prognostication using conditional survival estimates. Cancer. Oct 15 2013;119(20):3589–92. doi:10.1002/cncr.28273

18. Bonnett LJ, Tudur-Smith C, Williamson PR, Marson AG. Risk of recurrence after a first seizure and implications for driving: further analysis of the Multicentre study of early Epilepsy and Single Seizures. BMJ. Dec 7 2010;341:c6477. doi:10.1136/bmj.c6477

19. Brown JW, Lawn ND, Lee J, Dunne JW. When is it safe to return to driving following first-ever seizure? J Neurol Neurosurg Psychiatry. Jan 2015;86(1):60–4. doi:10.1136/jnnp-2013-307529

20. Schmedding E, Darde JB, Gappmeier B, et al. Epilepsy and driving in Europe. 2005. https://road-safety.transport.ec.europa.eu/system/files/2021-07/epilepsy_and_driving_in_europe_final_report_v2_en.pdf

21. Pitkanen A, Roivainen R, Lukasiuk K. Development of epilepsy after ischaemic stroke. Lancet Neurol. Feb 2016;15(2):185–197. doi:10.1016/S1474-4422(15)00248-3

22. Engel J, Jr. Concepts of epilepsy. Epilepsia. 1995;36 Suppl 1:S23-9. doi:10.1111/j.1528-1157.1995.tb01648.x

23. Chou CC, Shih YC, Chiu HH, et al. Strategic infarct location for post-stroke seizure. Neuroimage Clin. 2022;35:103069. doi:10.1016/j.nicl.2022.103069

24. Schaper F, Nordberg J, Cohen AL, et al. Mapping Lesion-Related Epilepsy to a Human Brain Network. JAMA Neurol. Sep 1 2023;80(9):891–902. doi:10.1001/jamaneurol.2023.1988

25. Troscher AR, Gruber J, Wagner JN, Bohm V, Wahl AS, von Oertzen TJ. Inflammation Mediated Epileptogenesis as Possible Mechanism Underlying Ischemic Post-stroke Epilepsy. Front Aging Neurosci. 2021;13:781174. doi:10.3389/fnagi.2021.781174

26. Ben-Ari Y, Dudek FE. Primary and secondary mechanisms of epileptogenesis in the temporal lobe: there is a before and an after. Epilepsy Curr. Sep 2010;10(5):118–25. doi:10.1111/j.1535-7511.2010.01376.x

27. Kadam SD, White AM, Staley KJ, Dudek FE. Continuous electroencephalographic monitoring with radio-telemetry in a rat model of perinatal hypoxia-ischemia reveals progressive post-stroke epilepsy. J Neurosci. Jan 6 2010;30(1):404–15. doi:10.1523/jneurosci.4093-09.2010

28. Shen Y, Gong Y, Ruan Y, Chen Z, Xu C. Secondary Epileptogenesis: Common to See, but Possible to Treat? Front Neurol. 2021;12:747372. doi:10.3389/fneur.2021.747372

29. Chen TS, Huang TH, Lai MC, Huang CW. The Role of Glutamate Receptors in Epilepsy. Biomedicines. Mar 4 2023;11(3)doi:10.3390/biomedicines11030783

30. Ferreira-Atuesta C, Dohler N, Erdelyi-Canavese B, et al. Seizures after Ischemic Stroke: A Matched Multicenter Study. Ann Neurol. Nov 2021;90(5):808–820. doi:10.1002/ana.26212

31. Doerrfuss JI, Holtkamp M, Vorderwulbecke BJ. The SeLECT 2.0 Score-Significance of Treatment With Antiseizure Medication. JAMA Neurol. Nov 1 2023;80(11):1252. doi:10.1001/jamaneurol.2023.3371

32. Schubert KM, Sinka L, Galovic M. The SeLECT 2.0 Score-Significance of Treatment With Antiseizure Medication-Reply. JAMA Neurol. Nov 1 2023;80(11):1252–1253. doi:10.1001/jamaneurol.2023.3374

33. Koepp MJ, Trinka E, Mah YH, et al. Antiepileptogenesis after stroke-trials and tribulations: Methodological challenges and recruitment results of a Phase II study with eslicarbazepine acetate. Epilepsia Open. Sep 2023;8(3):1190–1201. doi:10.1002/epi4.12735

34. Koepp M, Trinka E, Loscher W, Klein P. Prevention of epileptogenesis - are we there yet? Curr Opin Neurol. Feb 13 2024;doi:10.1097/WCO.0000000000001256

35. Labovitz DL, Hauser WA, Sacco RL. Prevalence and predictors of early seizure and status epilepticus after first stroke. Neurology. Jul 24 2001;57(2):200–6. doi:10.1212/wnl.57.2.200

36. Cordonnier C, Henon H, Derambure P, Pasquier F, Leys D. Influence of pre-existing dementia on the risk of post-stroke epileptic seizures. J Neurol Neurosurg Psychiatry. Dec 2005;76(12):1649–53. doi:10.1136/jnnp.2005.064535

