## Supplemental for "Association of the timing and type of acute symptomatic seizures with post-stroke epilepsy and mortality"

-- ONLINE SUPPLEMENT --

**Content**

### Supplemental Methods: Description of Cohorts

**Derivation cohorts**

The full cohort (n=4,552) consisted of nine international subcohorts participating in a registry assessing post-stroke seizures incepted as part of the SeLECT study. Four out of these subcohorts (Austria, Germany (2), Italy, Switzerland (1)) were part of the original SeLECT study, and five (Colombia, Germany (1), Portugal, Spain, Switzerland (2)) were included additionally.

**Austria**

In the Austrian cohort people with remote symptomatic seizures (RSyS) and controls (people without RSyS) were randomly selected from a larger cohort of consecutive people with a primary stroke diagnosis admitted to a tertiary referral center in Linz between Jan 1, 2005, and Dec 31, 2014.The study excluded people with transient ischemic attack (n=11), pre-existing brain lesions (i.e., intracranial tumor, trauma, or other; n=7), hemorrhagic stroke (n=97), history of seizures (n=8), cerebral venous thrombosis (n=5), death within days after stroke (n=9), and insufficient follow-up (n=48). Baseline and follow-up data were retrospectively extracted from medical records. 28 (6%) cases received endovascular thrombectomy either with a suction device (until 2011) or using a stent-retriever (from 2011 onwards). All participants included had face-to-face follow-up neurological interviews 3–6 months after stroke and then yearly. Follow-up had a median of 10 months (IQR 2–41). It was noted whether the participant had RSyS. If seizures were suspected, additional EEG and brain imaging (MRI or CT) were done. 459 participants were finally included.

**Colombia**

The initial Colombian retrospective cohortincluded patients aged 18 or older with diagnosis of acute ischemic stroke admitted to the stroke unit of a university hospital in Bogota between January 1, 2014, and December 31, 2019 (n=465). Stroke diagnosis was made by a neurologist and confirmed by neuroimaging. Patients with primary hemorrhagic stroke, transient ischemic attack, history of previous seizures and epileptogenic comorbidities (severe traumatic brain injury, brain tumors, history of neurological surgery and sinus vein thrombosis) were not included. Baseline characteristics and follow up information were extracted from medical records. Mean follow-up was 21 months and patients lost to follow-up were excluded (n=116). Additional exclusion criteria were applied for this study, ultimately including a total of 322 patients from the original cohort.

**Germany (1)**

The German case-control cohort (1) included participantswith a first-ever hemispheric stroke admitted to a tertiary referral center in Homburg between January 1, 2010, and December 31, 2016. Screened participants included those that were consecutively evaluated by the Neurology Department. Participants with seizures within the first 7 days after the onset of stroke symptoms were included. Controls were randomly selected among participants with stroke that were admitted during the same time but had no seizures within the first 7 days after stroke. Participants with TIA, primary cerebral hemorrhage or cerebral sinus/venous thrombosis were excluded, so that 182 were finally included. Baseline characteristics for cases and controls were independently extracted from medical records by two neurologists and cross-checked for accuracy and completeness. Follow-up was completed at 63 months (IQR 5–68) and data was obtained from retrospective medical chart reviews.

**Germany (2)**

The German cohort (2) included participants (final n=311) with a first-ever hemispheric stroke admitted to a tertiary referral center in Muenster between Jan 1, 2003, and March 31, 2010. The cohort excluded participants with recurrent stroke (n=225), only infratentorial stroke (n=322), hemorrhagic stroke (n=32), transient ischemic attack (n=64), or cerebral venous thrombosis (n=14). Participants who died in hospital (n=195), died before the interview (n=21), or were lost to follow-up (n=139), and participants declining participation (n=12) were also excluded. 311 participants were finally included. Baseline characteristics were extracted from medical records. Follow-up was completed after median 23 months (IQR 12–44) and all participants received a structured telephone interview.

**Italy**

The Italian cohort was part of a population-based study in the Udine district with 153 312 residents. The cohort included all patients with first-ever strokes occurring between April 1, 2007, and March 31, 2009. The cohort excluded participants with transient ischemic attacks (n=178), previous brain lesions (i.e., brain tumor; the exact number of participants not recorded), non-ischemic stroke (n=156), previous history of seizures or epilepsy (n=22), and missing time-to-event data (n=108), and participants who were deceased or lost to follow-up (n=94). Finally, 399 were included. A neurologist assessed participants within 48 h of admission. All participants were followed up by a face-to-face interview with study neurologists at 1, 6, and 24 months after the stroke.

**Portugal**

The Portuguese cohort included participants that were part of a prospective longitudinal study of consecutive adults with neuroimaging-confirmed anterior circulation ischemic stroke admitted to the stroke unit of a university hospital in Lisbon over a 24 months period. Those with past medical history of epileptic seizures, traumatic head injury requiring hospital admission, or brain surgery were excluded. All patients (final n=151) received standardized clinical and diagnostic assessment during admission and after discharge. A blinded phone interview and a clinical appointment was conducted at 6- and 12-months after stroke to access the occurrence of epileptic seizures and functional outcome.

**Spain**

The Spanish cohort included participants that were part of a multicenter prospective longitudinal study evaluating the development of epilepsy in adults aged 18 years or older with acute ischemic or hemorrhagic stroke admitted to one of six hospitals in Catalonia between August 2012 and November 2013. Patients were enrolled at hospital arrival by neurologists. Stroke diagnosis was performed by trained neurologists at each center and confirmed by neuroimaging. The initial study excluded people from whom signed informed consent was not possible to obtain (n=62), had more than 6 hours between symptom onset and emergency room arrival (n=77), tPA was administered previous to blood collection (n=11), was not possible to get blood samples (n=6), had no symptoms at hospital arrival (n=12), no definite diagnosis was made (n=32), or no sufficient blood sample was obtained (n=13). For inclusion in this study, further exclusion criteria was applied: past medical history of epilepsy (n=22), transient ischemic attack (n=101), subarachnoid hemorrhage (n=11), arteriovenous malformation (n=1), subdural hematoma (n=4), and insufficient data from medical reports (n=12). Finally, 512 participants were included. Baseline and follow-up data were collected by chart review and phone interviews. All the patients were clinically monitored during hospitalization, and seizures were clinically diagnosed by certified neurologists.

**Switzerland (1)**

The Swiss cohort (1) included consecutive people aged 18 years or older with acute first- ever neuroimaging confirmed ischemic stroke admitted to a tertiary referral center in St Gallen, between Jan 1, 2002, and Dec 31, 2008. The cohort excluded people with transient ischemic attacks (n=495), previous history of stroke (n=250), primary hemorrhagic stroke (n=94), previous history of seizures (n=43), re-infarction during follow-up (n=9), and potentially epileptogenic co-morbidities (alcohol or drug abuse [n=60]; intracranial tumors [n=28]; cerebral venous thrombosis [n=11]; history of severe traumatic brain injury [n=12]; history of brain surgery [n=4]; or other, including cerebral arterio-venous malformations, large cerebral aneurysms, cerebral vasculitis, hydrocephalus, and cerebral abnormalities of undetermined etiology [n=15]). 85 participants were lost to follow-up or died before follow-up was done. A neurologist analyzed baseline characteristic at admission and a diagnosis of stroke was confirmed at discharge. Brain scan analysis was done using the best available imaging modality (MRI in 954 [80%] of 1200 participants vs CT in 246 [21%]) at discharge. All participants (final n=1200) were followed up after a median of 28 months (IQR 21–47) with a structured telephone interview based on a validated questionnaire to detect seizures. In participants who did not have the capacity to do the questionnaire, close relatives, nursing staff, or their general practitioner were interviewed. Positive answers triggered a face-to-face neurological consultation and an electroencephalogram (EEG) to determine the epileptic nature of these episodes and to exclude seizure mimics. If the neurologist suspected a cause of epileptic seizures other than the index ischemic stroke, follow-up imaging was requested to rule out a co-pathology or re-infarction.

**Switzerland (2)**

The Swiss cohort (2) was part of the Biomarker Signature of Stroke Aetiology (BIOSIGNAL) study, a prospective, observational, multicenter, inception cohort study to evaluate and validate selected blood-biomarkers in patients with confirmed acute ischemic stroke. Participants older than 18 years who were admitted with ischemic stroke and gave informed consent were recruited at the University Hospital Zurich, Switzerland, between October 2014 and April 2022. Participants with transient ischemic attacks were not included. For the pooling of this study patients with potentially epileptogenic co-morbidities including intracranial tumors (n=17), history of severe traumatic brain injury (n=3); history of brain surgery (n=7), or other (cerebral arterio-venous malformations, large cerebral aneurysms, cerebral vasculitis, hydrocephalus, and cerebral abnormalities of undetermined etiology [n=25]) were excluded. There were no participants with a history of seizures before stroke or cerebral venous thrombosis. Three participants were lost to follow-up and 94 died before follow-up was done. Finally 1016 participants were included. The presence of acute symptomatic seizures within the first 7 days after stroke onset was noted in the case report form for each participant. All participants received standardized followed up after 3 and 12 months during an outpatient visit or via a structured telephone interview by a neurologist and it was noted whether they had any unprovoked remote symptomatic seizures. Long-term follow-up 60 months following stroke was performed using chart review.

**Validation cohorts**

For validation, only patients with ASyS were included from three cohorts:

**Japan**

The Japan cohort included consecutive individuals aged 18 years or older with acute, first-ever, neuroimaging-confirmed ischemic stroke who were admitted to the National Cerebral and Cardiovascular Center in Osaka within 3 days of symptom onset, between July 1, 2019, and December 31, 2023. Patients with a history of epileptic seizures, severe traumatic brain injury, intracranial tumors, cerebral venous thrombosis, or previous brain surgery were excluded. All patients (final n=1675, 19 with ASyS) underwent standardized clinical and diagnostic assessments during hospitalization, conducted by certified neurologists. The median follow-up duration was 11.7 months (IQR 3.3–24 months).

**Argentina:**

The Argentinian cohort included 691 participants from a retrospective observational study evaluating the development of post-stroke epilepsy (PSE) in adults aged 18 years or older with ischemic stroke, treated at a single institution between January 2012 and June 2020. 23 of this participants had ASyS. Stroke diagnosis was confirmed through neuroimaging, and patients were followed up for at least one year, which required at least two neurological check-ups—one within the first year and another after that period. Exclusion criteria included a past medical history of epilepsy, any potentially epileptogenic structural injury (such as subarachnoid hemorrhage, arteriovenous malformation, cerebral venous thrombosis, brain tumor, or severe head trauma), incomplete follow-up of less than a year, and missing MRI data. Data collection was done through medical chart reviews by four neurologists who documented the occurrence of unprovoked seizures, and an ad-hoc database was created. The dataset included demographic and cardiovascular risk factors, prior medical history, stroke classification using the TOAST system, stroke severity based on the NIH Stroke Scale (NIHSS), and functional disability measured by the modified Rankin Scale (mRS). MRI scans were analyzed for stroke topography and arterial territory, and white matter lesions were evaluated according to the Fazekas scale. Additional variables included family history of epilepsy, vascular risk factors (e.g., hypertension, diabetes, smoking), use of epileptogenic or antiseizure medications, and occurrence of acute symptomatic seizures during hospitalization. Post-discharge, seizure frequency was classified (e.g., daily, weekly, monthly), along with antiseizure medication use and electroencephalogram results.

**Switzerland (3)**

The Swiss cohort (3) involved a retrospective analysis of medical records from the Department of Epileptology and EEG at the University Hospital Zurich, Switzerland, between March 2002 and March 2024. Unlike the BIOSIGNAL study (Switzerland (2)), this cohort specifically focused on patients with ASyS (n=32) and RSyS who were not enrolled in the BIOSIGNAL study. A total of 32,157 patient records were screened for inclusion. The same inclusion and exclusion criteria as Switzerland (2) were applied, excluding patients with potentially epileptogenic comorbidities such as intracranial tumors, severe traumatic brain injury, or prior brain surgery. Participants with a history of seizures before stroke or cerebral venous thrombosis were not included. Follow-up duration was determined by the last epilepsy outpatient clinic visit, and standardized assessments were conducted to document the occurrence of ASyS and any subsequent unprovoked seizures. Detailed clinical and diagnostic data were collected, and long-term outcomes were evaluated through chart reviews to monitor seizure recurrence and stroke-related complications.

### Supplemental Methods: Informed consent procedures

All subjects in the Italian, Spanish, Swiss (2) and Portuguese cohort and those having a face-to-face interview in the Swiss (1) cohort gave written informed consent. All subjects evaluated by telephone in the Swiss (1) and German (2) cohorts gave verbal informed consent. According to Swiss and German law the regional ethical committees exempted these cohorts from requiring written informed consent. The Austrian and German (1) case-control studies were classified as retrospective service evaluation by the regional ethical committee and informed consent was not required. The retrospective Colombian cohort did not require informed consent by the regional ethical committee.

The use and analysis of patient data from the Zurich arm of the BIOSIGNAL study was done based on the project approval with BASEC- ID PB_2016-00672. that was approved by the Cantonal Ethics Commission of Zurich.

All other cohorts received approval from their local ethical committees.

### Supplemental Methods: Other definitions

We used definitions and classifications of the International League Against Epilepsy (ILAE) for seizures types and epilepsy ^1^ ^2^ ^3^ and the World Health Organization for stroke. ^4^

Stroke severity was measured with the National Institutes of Health Stroke Scale (NIHSS). Stroke etiology was classified according to the Trial of Org 10172 in Acute Stroke Treatment (TOAST) classification. ^5^

Supplemental Table 1: *Baseline characteristics of derivation cohorts (n=4552)*

| **Variable** | **N (%) or**  **median (IQR)** |
| --- | --- |
| Cohort |  |
| Austria | 459 (10%) |
| Colombia | 322 (7%) |
| Germany (1) | 182 (4%) |
| Germany (2) | 311 (7%) |
| Italy | 399 (9%) |
| Portugal | 151 (3%) |
| Spain | 512 (11%) |
| Switzerland (1) | 1200 (26%) |
| Switzerland (2) | 1016 (22%) |
| Age *(years)* | 73 (62-81) |
| Sex |  |
| Male | 2547 (56%) |
| Female | 2005 (44%) |
| NIHSS at admission |  |
| ≤3 | 1932 (42%) |
| 4-10 | 1545 (34%) |
| ≥11 | 1075 (24%) |
| Stroke location |  |
| Middle cerebral artery territory involvement | 3120 (69%) |
| Cortical involvement | 2332 (51%) |
| Stroke cause |  |
| Small-vessel occlusion | 893 (20%) |
| Large-artery atherosclerosis | 831 (18%) |
| Cardioembolism | 1374 (30%) |
| Other or undetermined | 1454 (32%) |
| Treatment |  |
| Acute reperfusion treatment | 1286 (28%) |
| ASM treatment started *(total cohort)* | 189 (4%) |
| ASM treatment after acute symptomatic seizure | 147/226 (65%) |
| ASM treatment after short acute symptomatic seizure | 126/182 (69%) |
| ASM treatment after acute symptomatic status epilepticus | 7/8 (88%) |
| Acute symptomatic seizure |  |
| Focal aware, short seizure | 58 (1.3%) |
| Focal with impaired awareness, short seizure | 36 (0.8%) |
| Focal to bilateral tonic clonic, short seizure | 88 (1.9%) |
| Status epilepticus | 8 (0.2%) |
| Undetermined | 36 (0.8%) |
| Duration of follow-up *(months)* | 18 (12-39) |

NIHSS, National Institutes of Healthy Stroke Scale; ASM, anti-seizure medication.

Supplemental Table 2: *Baseline characteristics according to early seizure days after ischemic stroke*

| **Variable** | **Day 0** | **Day ≥ 1** | **P-Value** |
| --- | --- | --- | --- |
| Total | 127 | 84 |  |
| Age *(years)* | 69 (53-85) | 73 (59-87) | 0.08 |
| Sex |  |  |  |
| Male | 77 (61%) | 48 (57%) | 0.62 |
| NIHSS at admission |  |  |  |
| ≤3 | 49 (39%) | 16 (19%) | **0.002** |
| 4-10 | 32 (25%) | 28 (33%) | 0.21 |
| ≥11 | 46 (36%) | 40 (48%) | 0.10 |
| Stroke location |  |  |  |
| Middle cerebral artery territory involvement | 92 (72%) | 68 (81%) | 0.15 |
| Cortical involvement | 79 (62%) | 54 (64%) | 0.76 |
| Stroke cause |  |  |  |
| Small-vessel occlusion | 4 (3.1%) | 3 (3.6%) | 0.86 |
| Large-artery atherosclerosis | 30 (24%) | 30 (35%) | 0.06 |
| Cardioembolism | 41 (32%) | 29 (35%) | 0.74 |
| Other or undetermined | 52 (41%) | 22 (26%) | **0.03** |
| Treatment |  |  |  |
| Acute reperfusion treatment | 38 (30%) | 36 (43%) | **0.05** |
| ASM treatment after ASyS | 76 (60%) | 62 (74%) | **0.03** |
| ASyS |  |  |  |
| Focal aware, short seizure | 28 (22%) | 27 (32%) | 0.11 |
| Focal with impaired awareness, short seizure | 18 (14%) | 13 (15%) | 0.79 |
| Focal to bilateral tonic clonic, short seizure | 57 (45%) | 27 (32%) | 0.06 |
| Status epilepticus | 1 (1%) | 5 (6%) | 0.06 |
| Undetermined | 18 (15%) | 10 (12%) | 0.63 |
| Duration of follow-up *(months)* | 12 (7-44) | 12 (4-38) | 0.22 |
| Remote symptomatic seizures (RSyS) | 35 (28%) | 19 (23%) | 0.42 |
| Death/ Mortality | 22/108 (17%) | 10/54 (12%) | 0.78 |

NIHSS, National Institutes of Healthy Stroke Scale; ASM, anti-seizure medication.

Data on the timing of the first ASyS was not available in 22 patients. ASyS were categorized into those occurring on day 0 and those after day 0 (Table 1). Significant differences were observed in stroke severity (lower on day 0), stroke cause (more undetermined cases and fewer cases with large artery atherosclerosis on day 0), acute reperfusion treatment (lower on day 0), ASM treatment (less frequent on day 0), and a tendency toward more generalized seizures on day 0, whereas status epilepticus occurred more frequently after day 0.

Supplemental Table 3: *Multivariable Cox regression model of time to first remote symptomatic seizure*

| **Variable** | **aHR (95% CI)** | **P value** |
| --- | --- | --- |
| Demographics |  |  |
| Age *(per 10 years)* | 0.9 (0.9-1.1) | 0.07 |
| Male sex | 1.0 (0.7-1.3) | 0.96 |
| Stroke severity at admission |  |  |
| NIHSS 4-10 | **1.8 (1.3-2.5)** | **0.001** |
| NIHSS ≥11 | **4.1 (2.9-5.7)** | **<0.001** |
| Stroke location |  |  |
| Middle cerebral artery territory involvement | **1.4 (1.1-2.0)** | **0.02** |
| Cortical involvement | **2.2 (1.7-2.9)** | **<0.001** |
| Stroke cause |  |  |
| Small-vessel occlusion | 1.0 (0.6-1.6) | 0.97 |
| Larger-artery atherosclerosis | **1.5 (1.1-2.1)** | **0.005** |
| Cardioembolic | 0.9 (0.7-1.2) | 0.53 |
| Treatment |  |  |
| Acute reperfusion treatment | 0.7 (0.6-1.0) | 0.07 |
| Acute symptomatic seizure type |  |  |
| Undetermined day | 2.6 (1.0-7.0) | 0.06 |
| FBTC-, day > 0 | **2.6 (1.4-5.0)** | **0.004** |
| FBTC+, day > 0 | **6.1 (2.8-13.0)** | **<0.001** |
| FBTC-, day 0 | **4.1 (2.4-7.0)** | **<0.001** |
| FBTC+, day 0 | **7.2 (4.5-11.6)** | **<0.001** |
| Status epilepticus | **12.7 (3.0-52.7)** | **<0.001** |

Data analyzed using a Cox proportional hazards model in the derivation cohort (n=4552). Dependent variable was time to death of any cause.

aHR, adjusted hazard ratio; NIHSS, National Institutes of Healthy Stroke Scale; ASM, anti-seizure medication.

Supplemental Table 4: *Results of different multivariate regression models of time to first remote symptomatic seizure in patients with acute symptomatic seizures.*

|  | **LASSO** | **Cox-regression** | | **Wald** | | **CRR** | |
| --- | --- | --- | --- | --- | --- | --- | --- |
| **Variable** | **Coefficients** | **aHR**  **(95% CI)** | **P value** | **aHR**  **(95% CI)** | **P value** | **aHR**  **(95% CI)** | **P value** |
| Demographics |  |  |  |  |  |  |  |
| Age *(per 10 years)* |  | 1.0 (1.0-1.0) | 0.32 |  |  |  |  |
| Stroke severity at admission |  |  |  |  |  |  |  |
| NIHSS 4-10 |  | 0.5 (0.2-1.1) | 0.06 |  |  | 0.6 (0.3-1.3) | 0.10 |
| NIHSS ≥11 |  | 0.7 (0.3-1.5) | 0.39 |  |  | 0.5 (0.2-1.1) | 0.10 |
| Stroke location |  |  |  |  |  |  |  |
| Middle cerebral artery territory involvement |  | 0.9 (0.5-1.8) | 0.82 |  |  | 1.0 (0.4-2.1) | 0.79 |
| Cortical involvement | **0.16** | 1.6 (0.9-3.0) | 0.11 | 1.6 (0.9-2.8) | 0.12 | 1.8 (0.9-3.7) | 0.06 |
| Stroke cause |  |  |  |  |  |  |  |
| Small-vessel occlusion |  | 1.4 (0.4-5.2) | 0.62 |  |  | 0.5 (0.0-5.6) | 0.55 |
| Large-artery atherosclerosis (LAA) |  |  |  |  |  |  |  |
| LAA, female |  | 2.9 (0.9-9.5) | 0.08 | 1.7 (0.6-4.8) | 0.08 | 2.9 (1.0-8.5) | **0.04** |
| LAA, male | **0.63** | 4.2 (1.6-10.9) | 0.003 | 2.9 (1.4-6.0) | **0.01** | 3.8 (1.7-8.6) | **0.003** |
| Cardioembolic |  | 2.0 (0.9-4.5) | 0.11 |  |  | 1.3 (0.6-3.0) | 0.46 |
| Treatment |  |  |  |  |  |  |  |
| Early ASM treatment |  | 1.0 (0.6-1.8) | 0.89 |  |  | 0.9 (0.4-1.9) | 0.94 |
| Acute reperfusion treatment |  | 1.6 (0.8-2.9) | 0.16 |  |  | 1.6 (0.7-4.1) | 0.30 |
| Acute symptomatic seizure type |  |  |  |  |  |  |  |
| FBTC-, day > 0 | **0.00** | 0.9 (0.3-3.1) | 0.9 | 1.0 (0.3-3.3) | 0.98 | 1.6 (0.2-2.4) | 0.49 |
| FBTC+, day > 0 |  | 1.2 (0.3-4.5) | 0.78 | 1.5 (0.4-5.2) | 0.51 | 1.4 (0.3-6.0) | 0.88 |
| FBTC-, day 0 |  | 1.2 (0.4-3.8) | 0.78 | 1.3 (0.4-4.0) | 0.64 | 1.1 (0.3-3.8) | 0.96 |
| FBTC+, day 0 | **0.52** | 2.1 (0.7-6.6) | 0.22 | 2.6 (0.9-7.9) | 0.09 | 2.6 (0.8-8.7) | 0.14 |
| Status epilepticus | **0.51** | 3.5 (0.8-15.9) | 0.11 | 4.3 (1.0-18.1) | **0.04** | 6.3 (1.1-36.4) | **0.04** |

Least Absolute Shrinkage and Selection Operator (LASSO) Cox-regression was utilized for variable selection and penalization to enhance prediction accuracy. The Wald backward regression tested the robustness of the LASSO model's predictions. Competitive risk regression (CRR), applying Fine and Gray’s subdistribution hazard model, considered the death as a competing event. Consistent variables emerged as predictive across models for the onset of post-stroke epilepsy.

Supplemental Table 5: *Multivariable Cox regression model of time to first remote symptomatic seizure in patients with acute symptomatic seizures.*

| **Variable** | **aHR** | **Assigned**  **integer value** | **P value** |
| --- | --- | --- | --- |
| Stroke location |  |  |  |
| Cortical involvement | 1.6 | 1 | 0.12 |
| Stroke etiology and sex |  |  |  |
| Large-artery atherosclerosis, female | 1.7 | 1 | 0.34 |
| Large-artery atherosclerosis, male | 2.9 | 2 | 0.005 |
| Acute symptomatic seizures type |  |  |  |
| FBTC-, day ≥ 1 | 1.0 | 0 | 0.98 |
| FBTC+, day ≥ 1 | 1.4 | 1 | 0.51 |
| FBTC-, day 0 | 1.2 | 1 | 0.64 |
| FBTC+, day 0 | 2.6 | 2 | 0.09 |
| Status epilepticus | 4.4 | 4 | 0.04 |

Data analysed using Wald backward-elimination of a Cox proportional hazards model in stroke survivors in patients with acute symptomatic seizures (n=233). Dependent variable was time to first remote symptomatic seizure after stroke. The integer value for all predictors was assigned by dividing their respective aHR by the aHR of the lowest three values and rounding to the nearest integer, as performed in the original SeLECT model development. Assigning integers to the Competitive risk regression (CRR), conducted via Fine and Gray’s subdistribution hazard model, adjusted for the presence of death as a competing risk but did not outpace the Wald model in predictive performance. As well combining both models was not superior.

aHR, adjusted hazard ratio; FBTC+, focal to bilateral tonic-clonic short seizure; FBTC-, other type of short seizure; day 0, same day as stroke onset.

Supplemental Table 6: *Cross-Validation using a leave-one-cohort-out strategy and calculating Coefficient of variance between centers:*

| **Select-ASyS** | **CoV 12 months** | **CoV 24 months** |
| --- | --- | --- |
| **0** | 0.22 | 0.22 |
| **1** | 0.18 | 0.17 |
| **2** | 0.13 | 0.12 |
| **3** | 0.10 | 0.08 |
| **4** | 0.08 | 0.06 |
| **5** | 0.08 | 0.07 |
| **6** | 0.09 | 0.07 |
| **7** | 0.08 | 0.06 |
| **mean** | 0.12 | 0.11 |
| **min** | 0.08 | 0.06 |
| **max** | 0.22 | 0.22 |

The Coefficient of Variance (CoV) was utilized as a statistical measure to assess the relative variability among subcohorts. Our approach employed a leave-one-cohort-out cross-validation technique. Initially, the subcohorts were divided into nine distinct datasets. In each iteration, one subcohort was excluded, and the predicted risk of unprovoked seizures at 12 and 24 months was calculated for the remaining subcohorts. This process was systematically repeated for each of the nine subcohorts, ensuring each was excluded once. The standard deviation relative to the mean of the nine predicted risks was then calculated to derive the overall CoV (CoV = standard deviation / mean). This procedure allowed us to ascertain the CoV at both 12 and 24 months for each SeLEC-ASyS score ranging from 0 to 7, subsequently leading to the derivation of mean, minimum, and maximum CoV values at these intervals.

Generally, CoV values below 15% are considered excellent, indicating strong consistency and reliability in data. Values between 15% and 25% are seen as tolerable, accounting for the inherent variability in medical research. In our study of Select-ASyS scores, we observed mean CoV values of 12% at 12 months and 11% at 24 months, demonstrating remarkable consistency across datasets, well within the excellent range.

Supplemental Table 7: *Characteristics of patients with acute symptomatic seizures in the validation cohorts (n=74) and their association with RSyS.*

| **Variable** | **N (%) or**  **median (IQR)** | **HR (95% CI)** | **P value** |
| --- | --- | --- | --- |
| Cohort |  |  |  |
| Switzerland (3) | 32 (43%) |  |  |
| Argentina | 23 (31%) |  |  |
| Japan | 19 (26%) |  |  |
| Demographics |  |  |  |
| Age *(per 10 years)* | 7.2 (5.8-7.9) | 1.0 (1.0-1.1) | 0.47 |
| Male sex | 54 (73%) | 1.0 (0.3-3.0) | 0.94 |
| Stroke severity at admission |  |  |  |
| NIHSS 4-10 | 25 (34%) | 0.7 (0.3-2.1) | 0.57 |
| NIHSS ≥11 | 25 (34%) | 2.6 (0.9-7.4) | 0.08 |
| Stroke location |  |  |  |
| Middle cerebral artery territory involvement | 28 (38%) | 2.1 (0.7-5.9) | 0.18 |
| Cortical involvement | 64 (87%) | 2.2 (0.3-16.5) | 0.45 |
| Stroke cause |  |  |  |
| Small-vessel occlusion | 1 (1.4%) | 0.0 (0.0->100) | 0.81 |
| Larger-artery atherosclerosis | 22 (30%) | 1.4 (0.5-3.7) | 0.54 |
| Cardioembolic | 26 (35%) | 1.4 (0.5-3.8) | 0.52 |
| Treatment |  |  |  |
| Acute reperfusion treatment | 28 (38%) | 0.9 (0.3-2.6) | 0.82 |
| ASM treatment after ASyS | 63 (85%) | 0.3 (0.1-0.8) | **0.02** |
| Acute symptomatic seizure type |  |  |  |
| FBTC-, day > 0 | 20 (27%) | 0.4 (0.1-1.6) | 0.19 |
| FBTC+, day > 0 | 29 (39%) | 1.0 (0.3-3.0) | 0.94 |
| FBTC-, day 0 | 5 (7%) | 1.5 (0.2-11.2) | 0.72 |
| FBTC+, day 0 | 18 (24%) | 1.7 (0.6-4.3 | 0.30 |
| Status epilepticus | 6 (8%) | 0.8 (0.1-6.1) | 0.82 |

Data analyzed using univariable Cox proportional hazards regression. Dependent variable was time to first remote symptomatic seizure after stroke.

HR, hazard ratio; NIHSS, National Institutes of Health Stroke Scale

Supplemental Figure 1: *Predicted Risk of Remote Symptomatic Seizures According to the Updated SeLECT-ASyS and original SeLECT score*

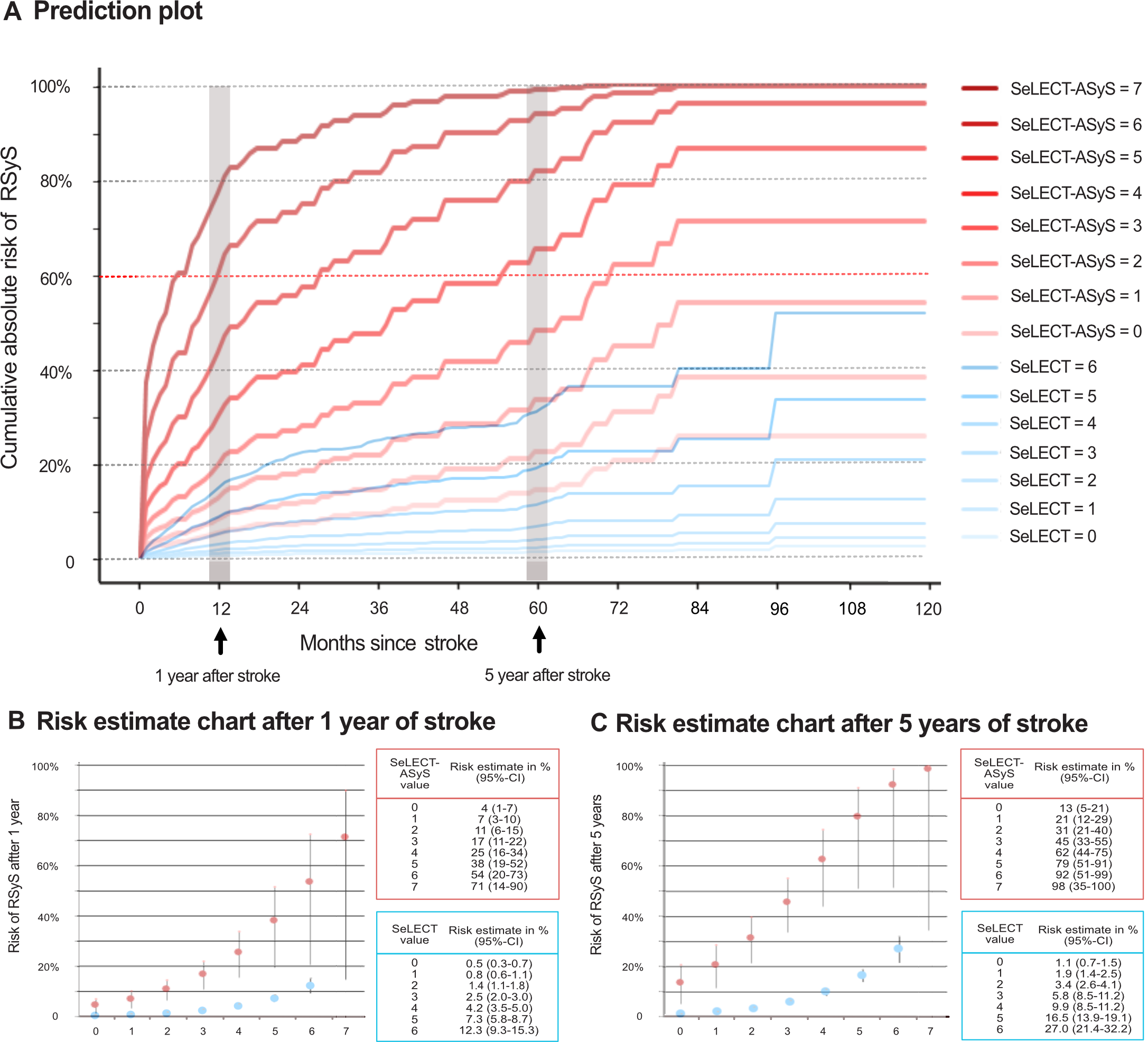

Panel A shows the predicted risk of unprovoked remote symptomatic seizures 0-120 months after stroke (in red SeLECT values for patients with ASyS and in blue for patients without ASyS). Each curve represents the estimates for a SeLECT-ASyS or original SeLECT_2.0_ value (curves with values ranging from 0-7 in red for patients with ASyS and with values ranging from 0-6 in blue for patients without ASyS). Risk estimate charts of remote symptomtic seizures (RSyS) 1 year and 5 years after stroke according to SeLECT-ASyS and SeLECT_2.0_ score are displayed in panels B and C, respectively. Vertical lines indicate 95% CIs.

Supplemental Figure 2: *Impact of different SeLECT_2.0_ score values and seizure-free intervals on the chance of an occurrence of a seizure in the next year*

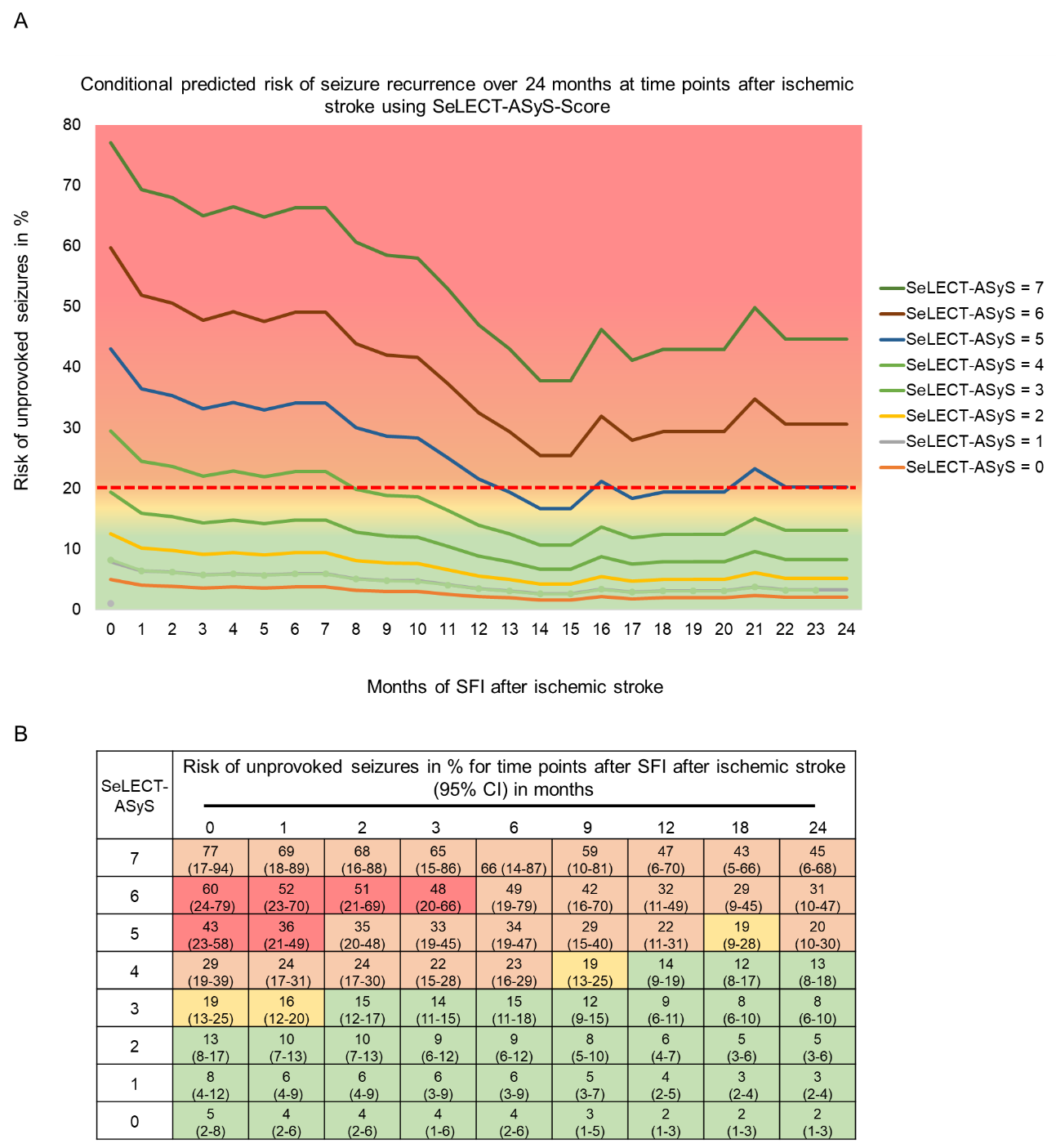

Panel A displays the impact of different seizure-free intervals (SFI) on the chance of an occurrence of a seizure in the next year (COSY) following ischemic stroke. The lines represent different SeLECT-ASyS scores. Panel B shows the numerical estimates of COSY stratified by different SFIs and SeLECT-ASyS values including the 95% confidence intervals. Colors suggested by different approaches by Bonnett et al.^6^ and Marson^7^ (acceptable range of risk for private driving for COSY of 20-40% suggested by Schmedding^8^): risk estimates ≥ 20% (and lower CI ≥ 20%) in red ("permissive approach"); risk estimates < 20% (and higher CI < 20%) in green ("conservative approach") in orange and yellow: orange when risk estimate ≥ 20% but lower CI < 20% (= "liberal approach"); yellow when risk estimate < 20% but upper CI > 20% (=" intermediate approach").

Supplemental Figure 3: *Timing of acute symptomatic seizure*

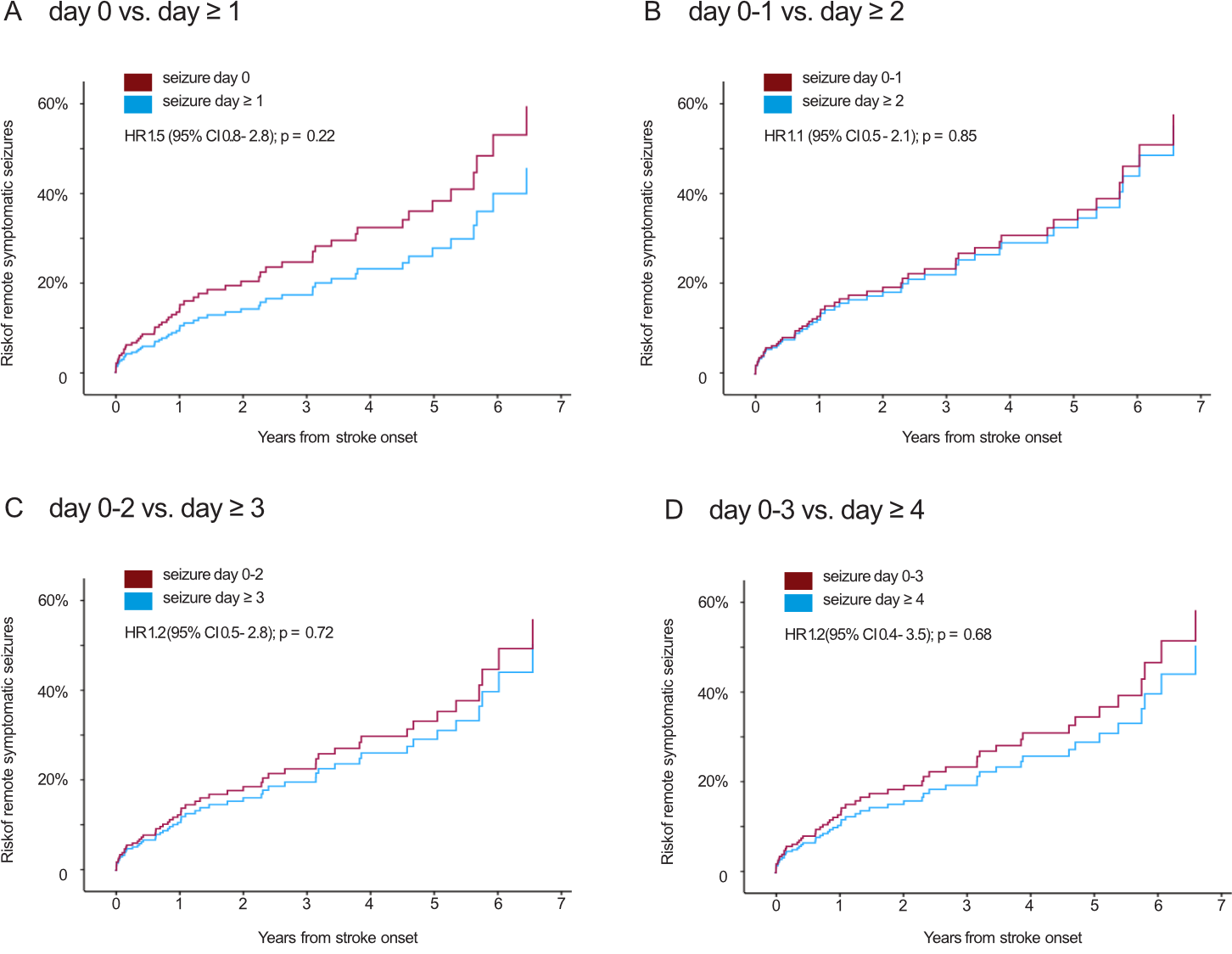
 Kaplan Meier estimates of the time to post-stroke epilepsy stratified by acute symptomatic seizure timing in patients with acute symptomatic seizures (n=211). All results were obtained after adjusting for covariates (age, sex, National Institutes of Health Stroke Scale score at admission, cortical involvement, involvement of the middle cerebral artery territory, stroke cause, reperfusion treatment, and antiseizure medication treatment after acute symptomatic seizure, seizure type).

Supplemental Figure 4: *Type of acute symptomatic seizure*

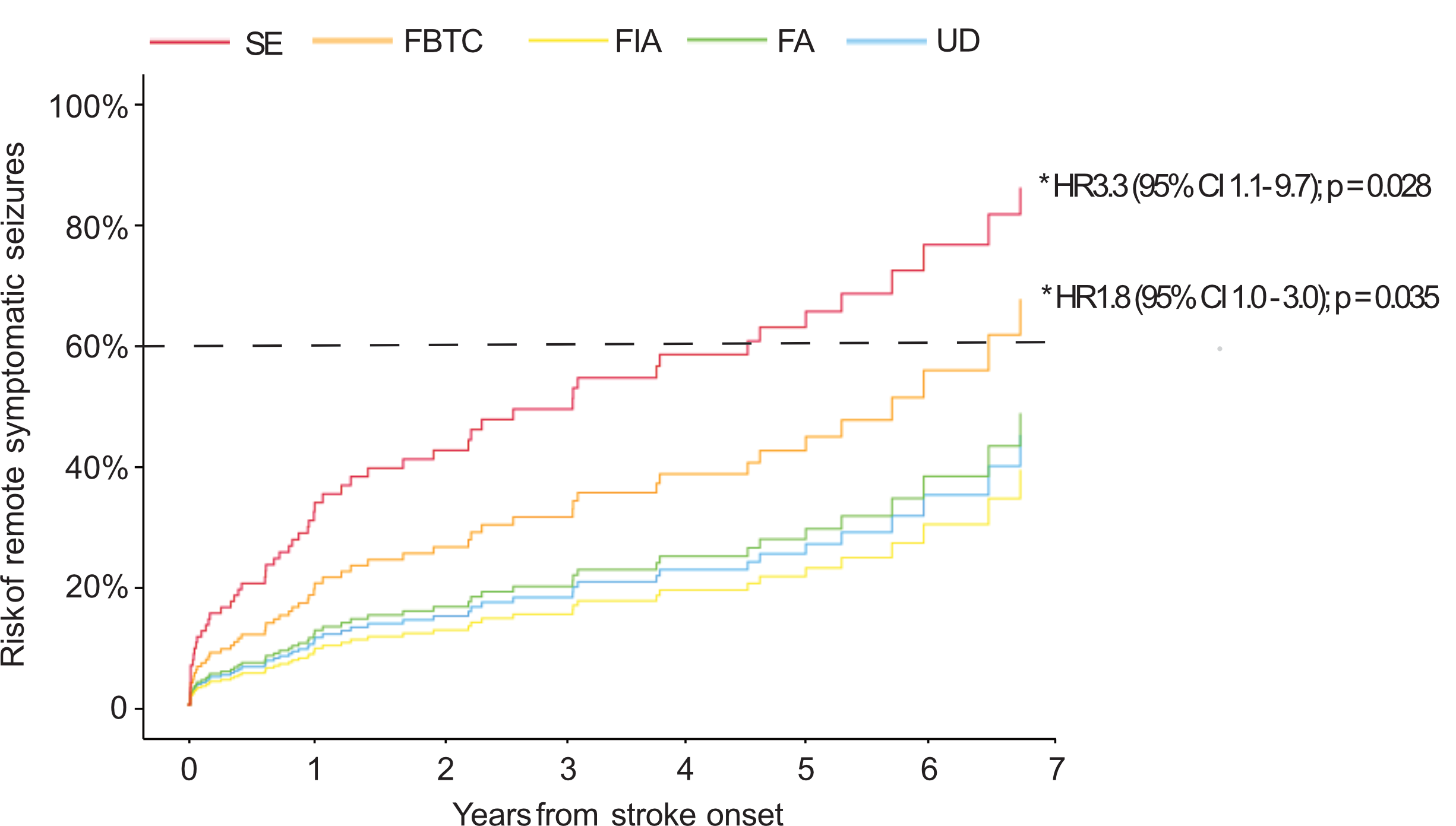

Kaplan Meier estimates of the time to post-stroke epilepsy stratified by acute symptomatic seizure type in patients with acute symptomatic seizures (n=226). All results were obtained after adjusting for covariates (age, sex, National Institutes of Health Stroke Scale score at admission, cortical involvement, involvement of the middle cerebral artery territory, stroke cause, reperfusion treatment, and antiseizure medication treatment after acute symptomatic seizure).

Among patients with acute symptomatic seizures, the highest risk for post-stroke epilepsy was observed in those with ASyS presenting as status epilepticus, followed by individuals with FBTCS, while other seizure types showed comparatively lower risk. Notably, both status epilepticus and FBTCS were independently associated with an increased likelihood of post-stroke epilepsy. In contrast, no significant associations were found for other seizure types in this particular cohort of patients experiencing acute symptomatic seizures.

Supplemental Figure 5: *Calibration plots of derivation cohorts*

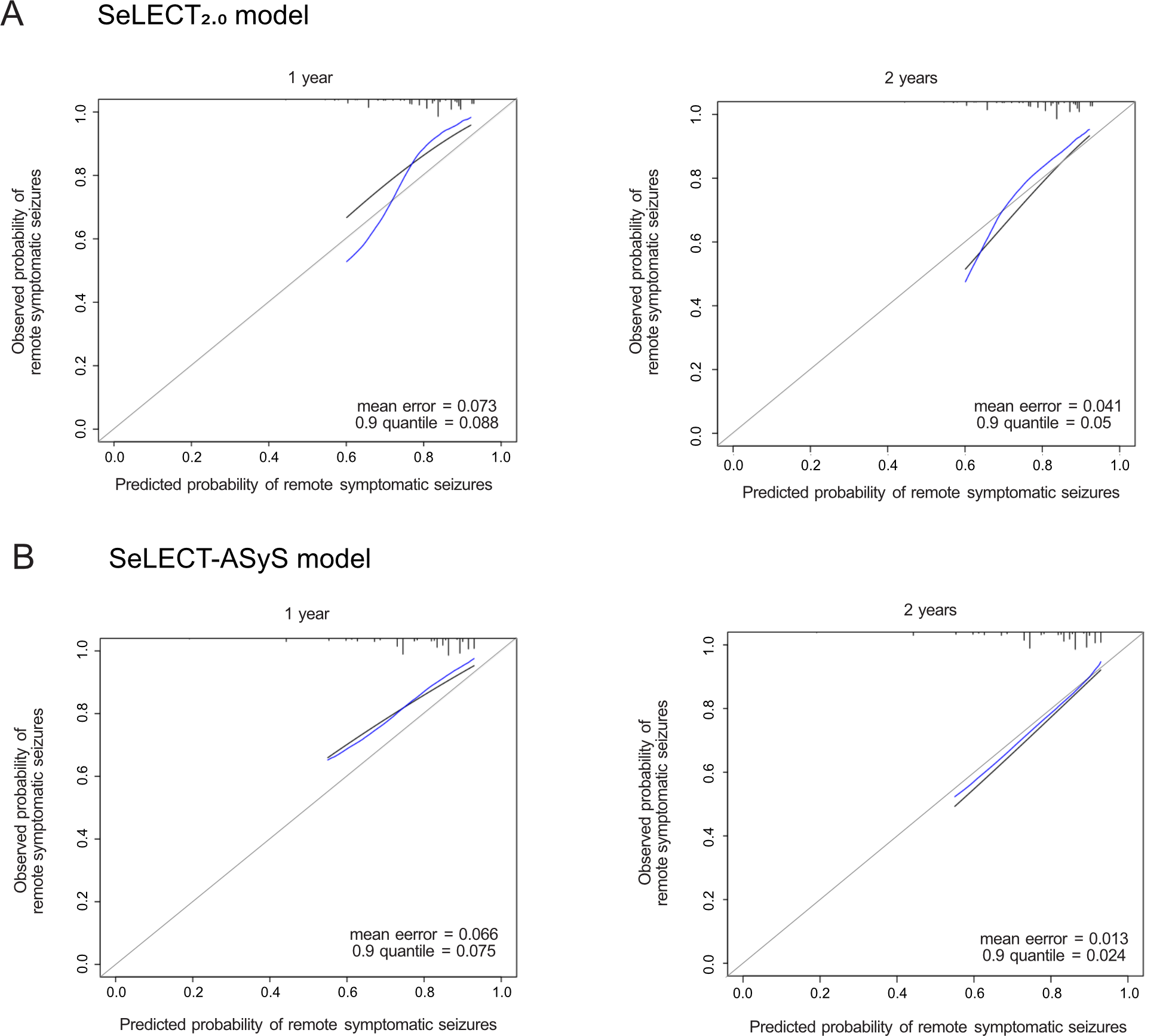

Calibration plots depicting the calibration performance of the previous SeLECT_2.0_ model (A) and the novel SeLECT-ASyS model (B) at 1-year (left) and 2-years (right) following stroke onset. The predicted outcomes are represented in black, optimism-corrected values (bootstrapped with 1000 samples) in blue, and the ideal calibration in gray. Notably, the SeLECT-ASyS model shows an improved calibration, especially with longer time of follow up.

Supplemental Figure 6: *C-statistics and calibration plots of the validation cohort*

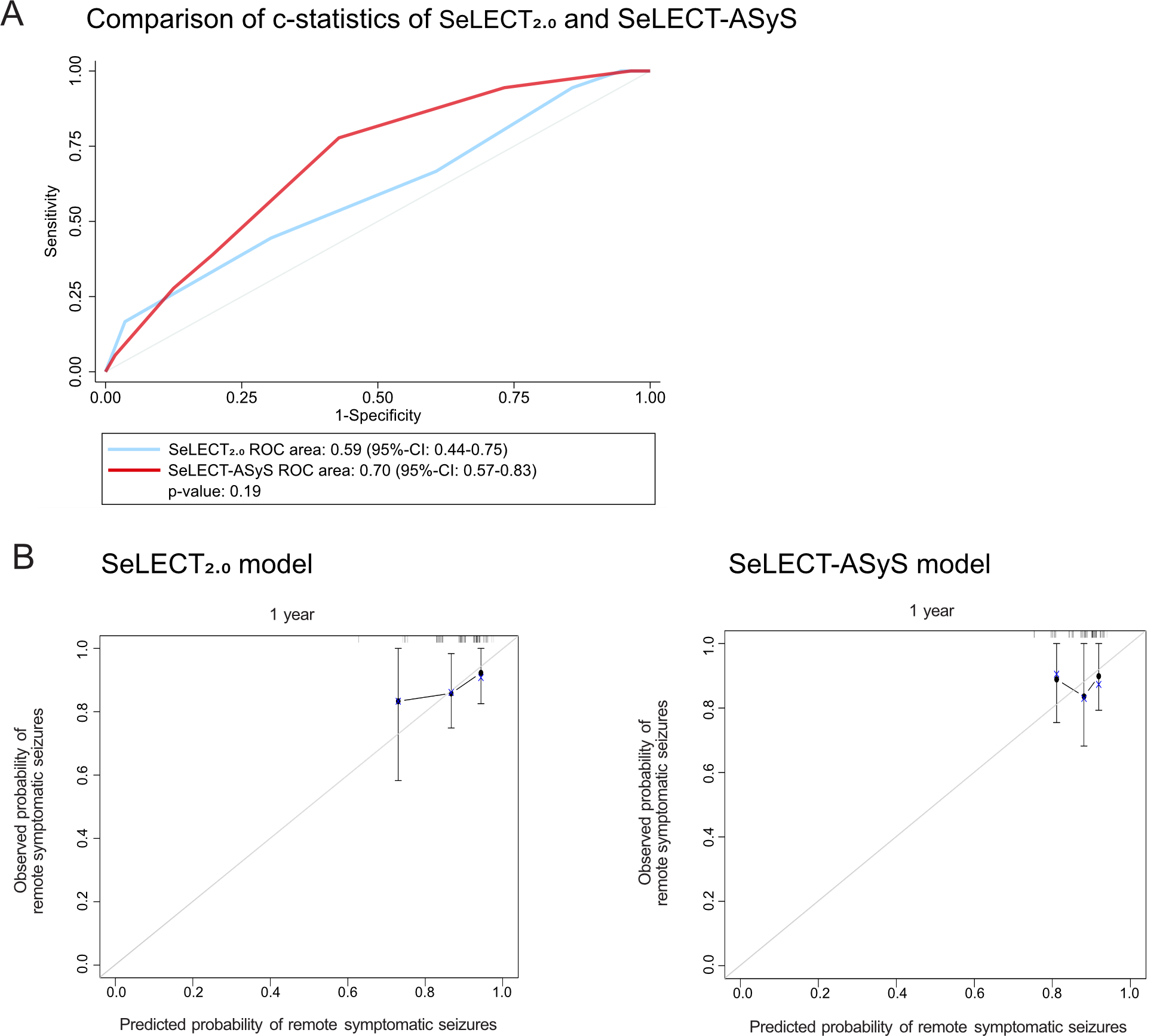

SeLECT-ASyS model demonstrated superior discrimination (Panel A) compared to the SeLECT_2.0_ model, with a C-statistic of 0.70 (95% CI: 0.57–0.83), indicating moderate predictive accuracy for remote symptomatic seizures. In contrast, the SeLECT 2.0 model showed a lower C-statistic of 0.59 (95% CI: 0.44–0.75). Both models exhibited good overall performance with low Brier scores of 0.11, indicating accurate probabilistic predictions for the risk of RSyS (Panels B).

**References:**

1.	Beghi E, D'Alessandro R, Beretta S, et al. Incidence and predictors of acute symptomatic seizures after stroke. *Neurology*. Nov 15 2011;77(20):1785-93. doi:10.1212/WNL.0b013e3182364878

2.	Fisher RS, Acevedo C, Arzimanoglou A, et al. ILAE official report: a practical clinical definition of epilepsy. *Epilepsia*. Apr 2014;55(4):475-82. doi:10.1111/epi.12550

3.	Fisher RS, Cross JH, French JA, et al. Operational classification of seizure types by the International League Against Epilepsy: Position Paper of the ILAE Commission for Classification and Terminology. *Epilepsia*. Apr 2017;58(4):522-530. doi:10.1111/epi.13670

4.	Aho K, Harmsen P, Hatano S, Marquardsen J, Smirnov VE, Strasser T. Cerebrovascular disease in the community: results of a WHO collaborative study. *Bull World Health Organ*. 1980;58(1):113-30.

5.	Adams HP, Jr., Bendixen BH, Kappelle LJ, et al. Classification of subtype of acute ischemic stroke. Definitions for use in a multicenter clinical trial. TOAST. Trial of Org 10172 in Acute Stroke Treatment. *Stroke*. Jan 1993;24(1):35-41. doi:10.1161/01.str.24.1.35

6.	Bonnett LJ, Tudur-Smith C, Williamson PR, Marson AG. Risk of recurrence after a first seizure and implications for driving: further analysis of the Multicentre study of early Epilepsy and Single Seizures. *BMJ*. Dec 7 2010;341:c6477. doi:10.1136/bmj.c6477

7.	Marson T. Evidence-based policy for driving: easier said than done. *Practical Neurology*. 2022;22(4):266-267. doi:10.1136/practneurol-2022-003458

8.	Schmedding E, Darde JB, Gappmeier B, et al. *Epilepsy and driving in Europe*. 2005. <https://road-safety.transport.ec.europa.eu/system/files/2021-07/epilepsy_and_driving_in_europe_final_report_v2_en.pdf>
